# Meta-analysis of genetic regulation of RNA editing in the human brain identifies new genes underlying neurological disease

**DOI:** 10.1101/2025.09.30.25337026

**Authors:** Winston H. Dredge, BP Kailash, Aline Réal, Benjamin Z. Muller, Beomjin Jang, Ting Fu, Hyomin Seo, Hong-Hee Won, Michael S. Breen, Xinshu Xiao, David A. Knowles, Jack Humphrey, Towfique Raj

## Abstract

Adenosine-to-inosine (A-to-I) RNA editing is among the most abundant post-transcriptional modifications, whose role in homeostasis and disease of the central nervous system (CNS) has been extensively reported. Regulation of A-to-I editing by *cis*-acting genetic variants (edQTLs) is understudied, especially in the brain. As a part of the BigBrain Project (n=4,471 individuals), which represents a >10-fold increase in sample size from previous edQTL studies, we harmonized A-to-I sites across datasets to perform a meta-analysis that identified 38,731 edQTLs in 3,554 genes. Integrating other QTL summary statistics from BigBrain, we show edQTLs in 3’ untranslated regions (3’UTR) are more likely to share a genetic effect with gene expression or splicing. Using colocalization analysis, we identify 24 GWAS loci for neurological disorders explained by edQTLs alone, or in concert with other molecular QTLs. This study represents a valuable resource to the field of RNA editing and neurogenetics.

## Introduction

Genome-wide association studies (GWAS) have identified thousands of genomic loci harboring variants associated with disease. Over 90% of GWAS single nucleotide polymorphisms (SNPs) overlap non-coding regions of the genome, where variants must be interpreted through the vast landscape of gene regulatory biology^1^. Molecular quantitative trait locus (QTL) analyses are a widely adopted tool for resolving the downstream effects of non-coding genetic variants. Large-scale studies have identified SNPs which regulate the expression (eQTL) of nearly all protein-coding genes, and demonstrated enrichment of eQTL SNPs in GWAS loci^2,3^. However, the biological underpinnings of most disease risk loci remain unclear, emphasizing the need to consider other consequential molecular processes, including RNA modifications^4^.

Over 170 types of RNA modifications have been identified to date^5^. Adenosine-to-inosine (A-to-I) RNA editing is one of the most abundant post-transcriptional modifications, with its role in homeostasis and disease of the central nervous system (CNS) having been extensively reported^6–14^. Adenosine Deaminases Acting on RNA (ADAR) enzymes, which recognize double-stranded RNA (dsRNA) substrates, catalyze the conversion of A-to-I at an estimated 100 million sites transcriptome-wide^15^. A-to-I editing acts as a critical regulator of innate immune responses, and accordingly it has been shown that *cis*-acting genetic variants which influence levels of A-to-I editing (*cis*-edQTL) explain a larger portion of GWAS heritability for autoimmune and inflammatory disorders than QTLs for expression (eQTL) or RNA splicing (sQTL)^16^. Furthermore, edQTL-mediated heritability for a given disease is enriched in the relevant tissue of origin for that disease, such as cardiovascular tissues for coronary artery disease, or brain tissues for neurodegenerative and neuropsychiatric diseases^7,16^.

To maximize discovery power for edQTLs in the brain, we collected and uniformly processed RNA-sequencing (RNA-seq) and genotype data from 11 independent cohorts with CNS tissues as part of the BigBrain Project. Through rigorous harmonization, we prioritized high-confidence A-to-I editing sites, which were systematically annotated with cellular specificity, and prioritized for validation of their effect on RNA abundance by massively parallel reporter assay (MPRA). We performed a multi-ancestry meta-analysis of *cis*-edQTLs using a linear mixed model^17^. We leveraged colocalization analyses to test whether edQTLs share a genetic basis with GWAS loci for neurodegenerative and psychiatric disorders.

## Results

### Quantification and harmonization of A-to-I RNA editing in the BigBrain Project

The BigBrain Project consists of 10,202 RNA-seq samples from 4,471 unique donors from 11 independent studies sourced from multiple regions of the brain and spinal cord (**Fig. 1; Supplementary Note**). Following uniform processing of RNA-sequencing data, editing site calling was performed independently by tissue and cohort (tissue-cohort pairs, hereby referred to as “datasets”), to maximize A-to-I site discovery and reduce noise contributed by factors including sequencing batch and differences in study design (**Table 1**). Using *de novo* calling in combination with a supervised approach^18,19^, we identified a total of 706,560 high-confidence A-to-I sites across the 43 datasets (**Fig. 2a**).

**Figure 1.**
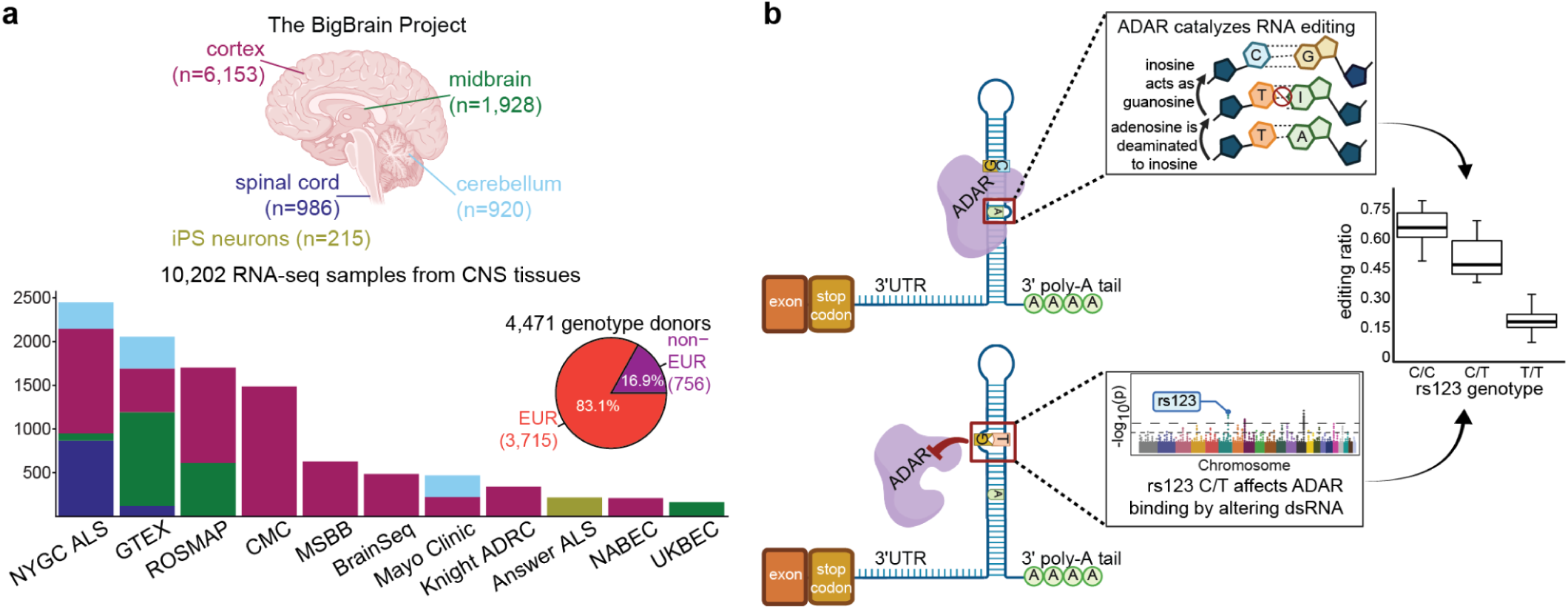
RNA editing in the BigBrain Project. **a)** Samples were sourced from various regions of the central nervous system (CNS) and induced pluripotent stem cell (iPS)-derived neurons, and are colored by broad tissue categories. Raw transcriptomic and genomic data was collected for a total n=10,202 RNA-seq samples from n=4,471 distinct individuals, of which 756 are of non-European (non-EUR) genetic ancestry. The 11 independent studies which make up BigBrain are: the New York Genome Center ALS Consortium (NYGC ALS), the Adult Genotype Tissue Expression project v8 (GTEX), the Religious Orders Study and Memory and Aging Project (ROSMAP), the CommonMind Consortium (CMC), the Mount Sinai/JJ Peters VA Medical Center Brain Bank (MSBB), the BrainSeq Consortium, the Mayo Clinic, the Knight Alzheimer’s Disease Research Center (Knight ADRC), the Answer ALS research study (Answer ALS), the North American Brain Expression Consortium (NABEC), and the United Kingdom Brain Expression Consortium (UKBEC). **b)** A cis-edQTL is an association between editing level measured for a given editing site and the genotype of a SNP within a +/- 1Mb window. In this schematic, the “C” allele (top panel, in blue) for SNP rs123 is associated with higher RNA editing (see boxplot panel), whereas the “T” allele (bottom panel, orange) causes a bulge in the double-stranded RNA (dsRNA) substrate, preventing efficient binding by ADAR, thus it is associated with a reduced level of editing.

**Figure 2.**
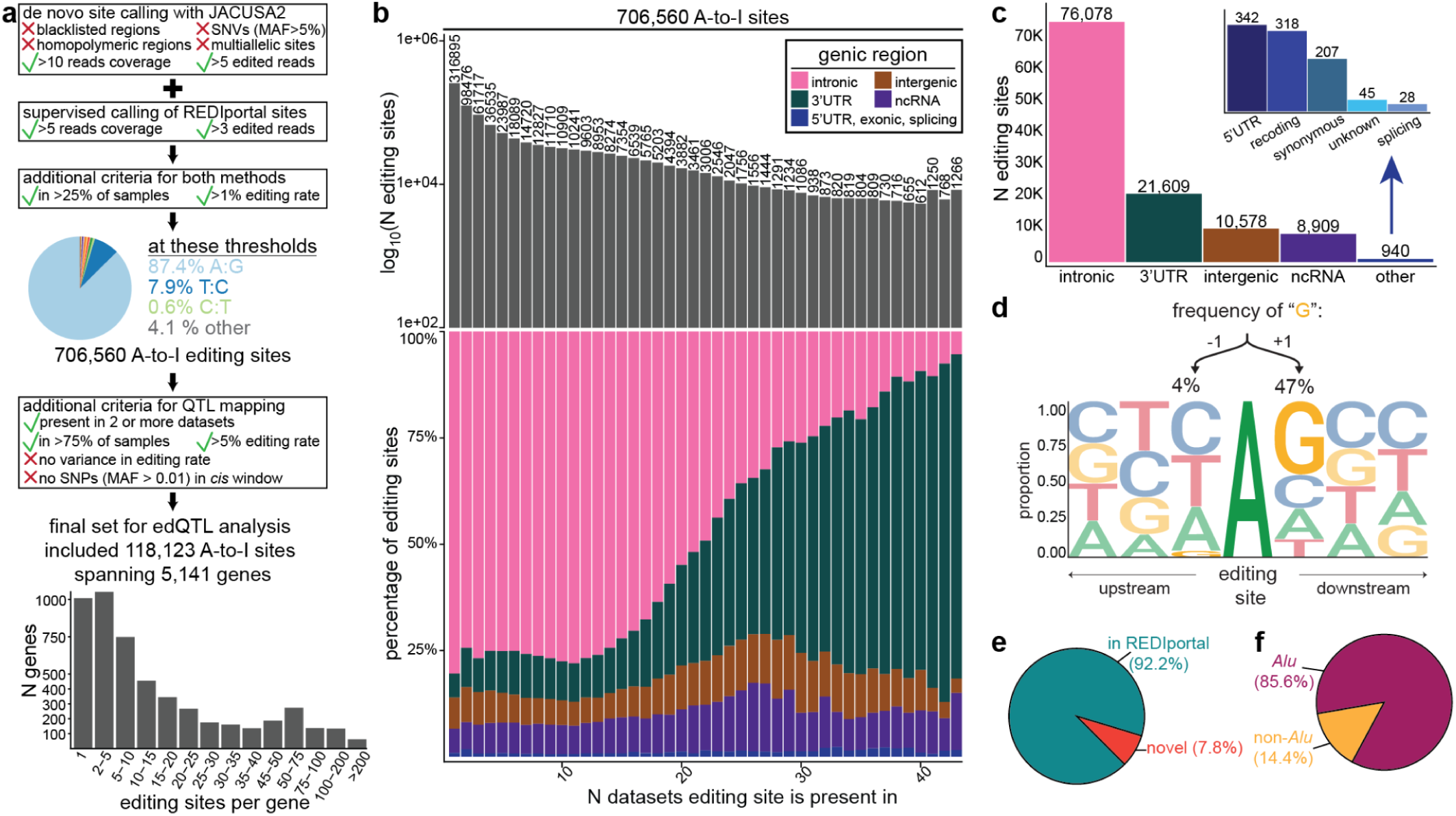
Quantifying RNA editing in 43 datasets. **a)** Analysis pipeline for calling RNA editing events from RNA-seq. We combined de novo identification of A-to-I sites and supervised calling of A-to-I sites which are documented in the REDIportal database. Sites were filtered based on editing rate, read coverage, replication across samples, and removed when overlapping blacklisted genomic regions or common genetic variants, defined by a minor allele frequency (MAF) > 5%. Following harmonization across datasets, a total of 118,123 high-confidence A-to-I sites in 5,141 genes were identified, with the distribution of the number of sites per gene shown. **b)** Prior to harmonization, 706,560 A-to-I sites were identified. The distribution of editing sites by the number of datasets they were detected in (x-axis) is represented by grey bars on a logarithmic scale (y-axis) with N displayed above each bar. Below each grey bar, the proportion of those N sites which map to various genomic regions, namely: intron (pink); 3’ untranslated region (3’UTR, green); intergenic (+/- 10 Kb, brown); non-coding RNA (ncRNA, purple); 5’ UTR, exon, splice sites (blue). **c)** Breakdown of the gene regions the 118,123 A-to-I sites map to. **d)** Sequence motif plot illustrating the proportion of cytosine (C, blue), guanosine (G, yellow), thymine (T, red), and adenosine (A, green) at +/- 3 bp flanking the edited “A”. Proportion of editing sites which were **e)** previously documented in REDIportal (teal) versus novel in BigBrain (orange) or **f)** reside within Alu repeats (magenta) versus other repetitive elements (yellow).

Approximately 45% of A-to-I sites were only detectable in one dataset, and only 1,286 sites replicated across all 43 datasets (**Fig. 2b**). Given that editing site discovery was highly variable between datasets, and not purely explained by sample size (*R* = 0.29; two-sided Pearson correlation test), we investigated correlation with various technical and biological variables within each dataset and across all datasets (**Supplementary Fig. 1**). Read length (*R* = 0.53) and intronic base coverage (*R* = 0.44) were the strongest positive predictors of editing site discovery (**Supplementary Fig. 1**). The ubiquitously expressed *ADAR* was significantly positively correlated with editing site discovery in 35/43 datasets compared to 38/43 datasets for the neural specific isoform, *ADARB1* (**Supplementary Fig. 1**). It is well established that the majority of ADAR-mediated RNA editing occurs in introns, but these regions can be particularly challenging to compare between datasets due to low and/or variable read coverage. Accordingly, when incrementally increasing the requirement of an A-to-I site to replicate in more datasets, there was a depletion in the proportion of intronic sites and an enrichment in sites in 3’ UTRs, where read coverage is more consistent (**Fig. 2b**).

In light of the high variation in editing sites between datasets, we further refined our catalog based on considerations for meta-analysis, namely editing sites were required to: have a mean editing rate > 5%, replicate in 2 or more datasets, and be present in > 75% of samples within each dataset. Editing sites with no variance were also removed, along with those which had no SNPs (minor allele frequency, MAF > 0.01) in a ±1 Mb window, leaving 118,123 A-to-I sites for our edQTL meta-analysis (**Fig. 2a**; **Table 2**). These editing events spanned 5,141 genes, with the vast majority of genes harboring a small number of editing events (**Fig. 2a**). However, some genes harbor thousands of A-to-I sites, such as *KCNIP4*, which encodes a voltage-gated potassium channel and has previously been shown to harbor intronic hyper-editing hotspots^19^. Most A-to-I sites in this final set resided in introns or 3’ UTRs, with only 570 sites in coding exons (**Fig. 2c**). These exonic editing events were spread across 190 genes, with 318 sites considered “recoding” events, which change the amino acid sequence encoded by the RNA molecule (**Table 3**)^20^.

The presence of both A-to-I and U-to-C edits resulting from A-to-I editing on overlapping sense and antisense transcripts has been documented^16,21^. Here, to uniformly call editing, all datasets were treated as unstranded RNA-seq, so we cannot directly link editing events to their strand of origin. To investigate how many T:C conversions from the final set of editing sites represent such events, we asked how many of these sites overlap multiple GENCODE transcripts. In addition to the many instances of overlapping genes genome-wide, longer 3’ UTR usage is highly abundant in the nervous system, and historically poorly annotated in reference transcriptomes^22,23^. To additionally capture instances where unannotated 3’ UTRs may overlap, we extended the 3’ UTR sequence for all GENCODE transcripts by 10 Kb. While T:C conversions represent only 7.3% of all editing sites, they represent 27.7% of sites overlapping inversely oriented genes (**Supplementary Fig. 2**). ANNOVAR^24^ is implemented in our analysis pipeline to map editing sites to genes, but in the case of these potentially ambiguous annotations, we report all overlapping genes (**Table 4**).

Previously, fluorescence-activated nuclei sorting (FANS) of human brain tissues was used to define editing sites that are specific to oligodendrocytes, glutamatergic neurons, and GABAergic neurons^19^. Here, we augment this published resource by harmonizing it with an independent FANS study^25^ to additionally define editing sites specific to microglia and astrocytes (**Supplementary Fig. 3**). From the set of A-to-I sites selected for meta-analysis, approximately 17.5% could be annotated with cellular resolution (**Supplementary Fig. 3**). We identified 7,293 sites specific to glial cells, and 13,325 sites specific to neurons (**Supplementary Fig. 3**).

In the genomic sequence flanking these A-to-I sites, we observed the expected depletion of guanosine +1bp upstream and enrichment of guanosine -1bp downstream of the edited adenosine (**Fig. 2d**). The final refined set of A-to-I sites were predominantly documented in the REDIportal database^26^ and reside within *Alu* elements (**Fig. 2f-g**). Altogether, this evidence suggests we are detecting *bona fide* ADAR-mediated editing events.

### Multi-ancestry RNA edQTL meta-analysis

While combining multiple independent studies into a single aggregate analysis by performing a meta-analysis can enable the identification of genetic associations with very small effect sizes, it is critical to evaluate heterogeneity between datasets to determine whether meta-analysis is suitable. Thus, prior to meta-analysis we verified that there are patterns of concordance in the effect sizes of edQTLs mapped in each BigBrain dataset (**Supplementary Fig. 4a**). Effect sizes were strongly correlated (*R* = 0.97) between datasets including those from different tissues or independent cohorts (**Supplementary Fig. 4b-d**).

Considering 17% of BigBrain donors are of non-European genetic ancestry, and several cohorts such as GTEx and the NYGC ALS consortium (*Humphrey et al., in preparation*) have multiple tissues sampled from the same individual, we leveraged the tool mmQTL for meta-analysis to implement a linear mixed model^17^. To break down the components of an edQTL association and demonstrate the utility of meta-analysis, we provide the example of an editing site in the 3’UTR of *VPS41*. This gene encodes a vacuolar sorting protein, and hypo-editing of this site has been previously observed in brain tissue from individuals with schizophrenia^7^. Here, we identify a significant edQTL association for this site, where the lead SNP resides 1,282 bp upstream (**Fig. 3a**).

**Figure 3.**
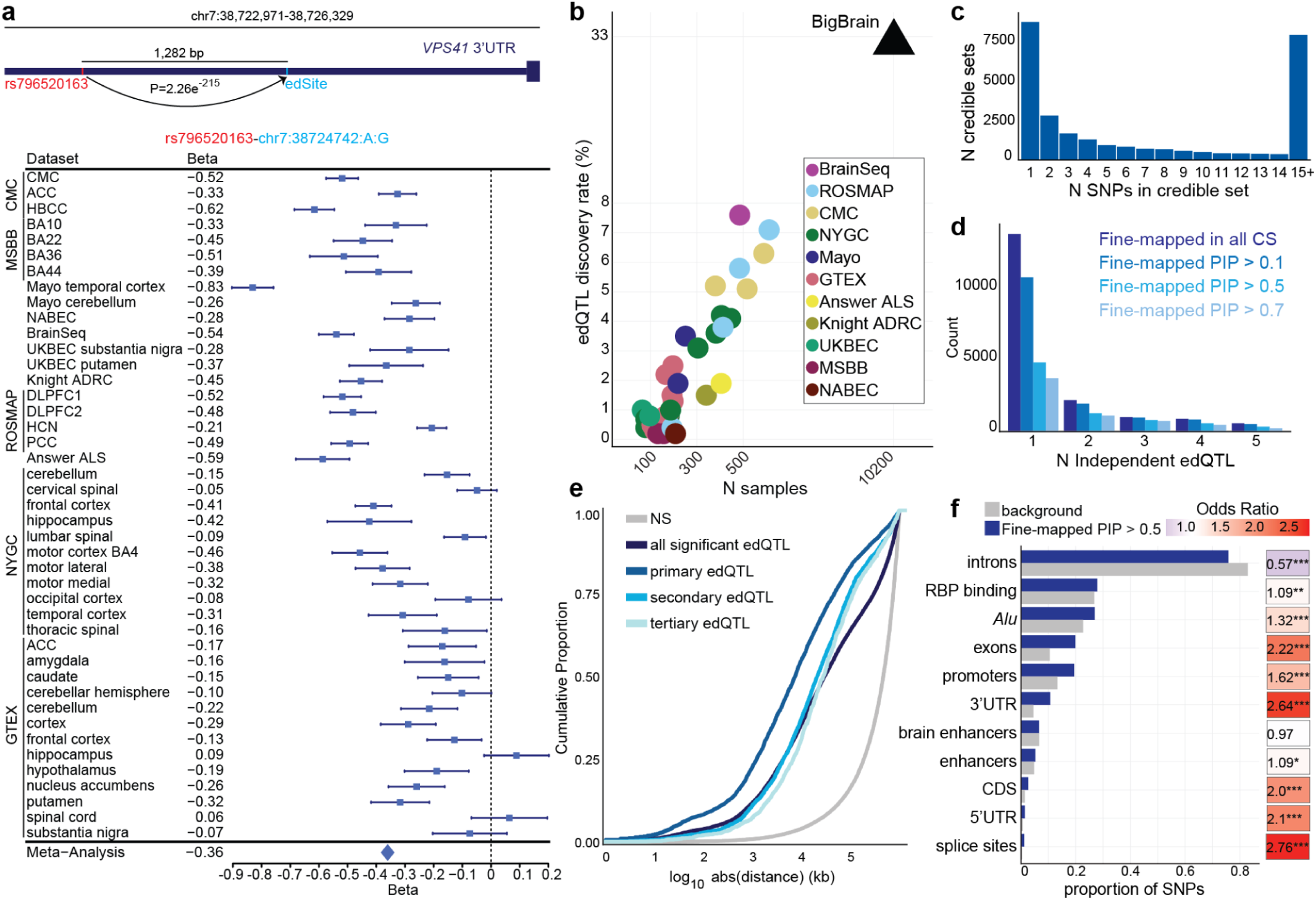
Meta-analysis powers large-scale RNA editing QTL discovery. **a)** Depiction of an edQTL in the 3’UTR of the negative-strand gene VPS41. The edQTL is for an editing site at chr7:38724742 (hg38, position in gene shown as blue dash), where the lead SNP rs796520163 (position in gene shown as red dash) is 1,282 bp upstream. The forest plot depicts the effect size (light blue square) with standard error bars (dark blue line) across all 43 BigBrain datasets, with a vertical black dashed line at 0. The meta-analysis random effect size and standard error are represented by a light blue diamond. **b)** edQTL discovery rate (y-axis) defined as the number of significant (5% FDR) edQTLs divided by the total number of edQTLs tested per dataset (tissue-cohort pair) against dataset sample size (x-axis). The black triangle represents the meta-analysis discovery rate, while circular points correspond to datasets (colored by cohort). The number of **c)** SNPs in fine-mapped edQTL credible sets and **d)** independent credible sets per edQTL signal. **e)** Cumulative distribution of distance from editing site to SNP for fine-mapped edQTLs, all significant edQTLs, and non-significant associations. **f**) Proportion of edQTL SNPs which could (PIP > 0.5, blue) or could not be fine-mapped (PIP < 0.1) overlapping various gene regions with enrichment analysis by Fisher’s exact test (P < 0.5 *, < 0.01 **, < 0.001*** ).

BigBrain represents an approximately 4-fold increase in edQTL discovery compared to previous work done in GTEx^16^, highlighting the power gained by meta-analysis. Amongst individual datasets, BrainSeq had the highest edQTL discovery rate at 7.6% (**Fig. 3b**). Meta-analysis dramatically improves edQTL discovery, identifying 38,731 edQTLs in 3,554 genes (33% discovery rate, FDR 5%) (**Fig. 3b**). Of all genes tested (n = 5,141) which had at least one A-to-I site, 69% had a significant edQTL. We took several measures to scrutinize the robustness of BigBrain edQTLs. To investigate the influence of one of the largest dataset, dorsolateral prefrontal cortex samples from ROSMAP, we excluded this dataset in a “hold-one-out” meta-analysis. The held-out meta-analysis replicated 35,544 edQTLs and 96% showed concordant direction of effect. A common approach for testing p-value replication is taking the non-null tests from a “discovery” dataset and estimating Storey’s π_1_ for non-null genes in a “replication” dataset^27^. However, this approach produces an asymmetric result based on the definition of “discovery” and “replication”, especially in the case where there is a significant discrepancy in sample size. Thus, in addition to Storey’s π_1_ we leveraged the pisquared method, which jointly fits a model of null and non-null p-values in both datasets, reporting a Jaccard similarity statistic^28^. By both measures, we observe strong replication of p-values in the held-out (Jaccard similarity index, π^2^ = 0.96; Storey’s π_1_= 0.74). Furthermore, we observed consistent replication (Storey’s π_1_ = 0.88 - 1.00) of previously published edQTLs across GTEx tissues in BigBrain (**Supplementary Fig. 4**).

Although mmQTL controls for population structure, enabling a multi-ancestry meta-analysis, we restricted downstream analyses to individuals of European ancestry (EUR, n=3,715) to support fine-mapping with an appropriate LD reference panel and comparison to GWAS which are limited in diverse ancestries. Our EUR-only meta-analysis revealed 37,506 edQTLs in 3,498 genes.

We applied statistical fine-mapping to infer likely causal variants for significant edQTLs, where 18,637 edQTLs (48%) could be assigned a 95% credible set (CS) (**Fig. 3c**). Of fine-mapped edQTLs, 82% had a SNP with a posterior inclusion probability (PIP) exceeding 0.1. Fine-mapping resolved 8,614 edQTLs to a credible set of 1 likely causal variant (**Fig. 3c**). For some editing sites, as many as five independent effects (i.e. five credible sets) were resolved (**Fig. 3d**). Previous edQTL studies have tested SNPs in a *cis*-window of ±100-200 Kb surrounding the editing site^7,16,29^. Here, to facilitate downstream statistical comparison of edQTL with QTLs for gene expression (eQTL) and splicing (sQTL) also mapped in BigBrain, as well as GWAS loci, we opted to test SNPs within a ±1 Mb window as standardly applied for *cis*-genetic associations. We observe enrichment of SNPs proximal to their editing sites, most pronounced in primary edQTLs which have a median distance of 7,398 bp to their respective editing site, as compared to a median distance of 29,879 bp for all significant edQTLs (**Fig. 3f**). Fine-mapped edQTL SNPs were depleted in introns (OR = 0.57, Fisher’s exact test), but strongly enriched in splice sites (OR = 2.76) and 3’ UTRs (OR = 2.64), as well as moderately enriched in *Alu* elements (OR = 1.3, **Fig. 3g**).

### edQTLs can be genetically linked to expression and splicing

We compared our significant edQTLs with eQTLs and sQTLs also mapped in BigBrain^30^. Of the 3,989 genes linked to an edQTL (including editing sites overlapping multiple genes), 3,790 (95%) of them were also significant eQTLs or sQTLs, with 2,305 genes (57.8%) being significant for all three QTL types (**Fig. 4a**). However, this overlap approach does not test whether different QTL types are caused by the same genetic variant. To test this, we performed colocalization using COLOC^31^ between all pairs of edQTLs and eQTLs, and edQTLs and sQTLs. COLOC calculates posterior probabilities (PP) for whether two phenotypes share the same causal SNP (PP4) or two independent causal SNPs (PP3), among other possibilities. At a PP4 > 0.8, we identified 602 (15%) edQTL genes with a shared eQTL, and 1296 (33%) with a shared sQTL (**Fig. 4b**), whereas at PP3 > 0.8 we found 520 independent eQTL genes and 135 independent sQTL genes. Taking the fine-mapping CS for the colocalized and independent molecular phenotype pairs where both QTL types could be fine-mapped, we observed that the colocalized pairs shared fine-mapped SNPs most of the time, whereas independent pairs rarely if ever shared SNPs (**Fig. 4c).** The precise location of an edQTL editing site (edSite) within a gene can reflect its role in mediating gene expression or splicing. While the majority of significant edQTL edSites are found within introns, we observed an enrichment for colocalized edQTL edSites within 3’UTR regions (P < 1e-16, Fisher’s exact test; **Fig. 4d**), suggesting RNA editing within 3’UTRs is more likely to have a functional connection to gene expression and/or splicing.

**Figure 4.**
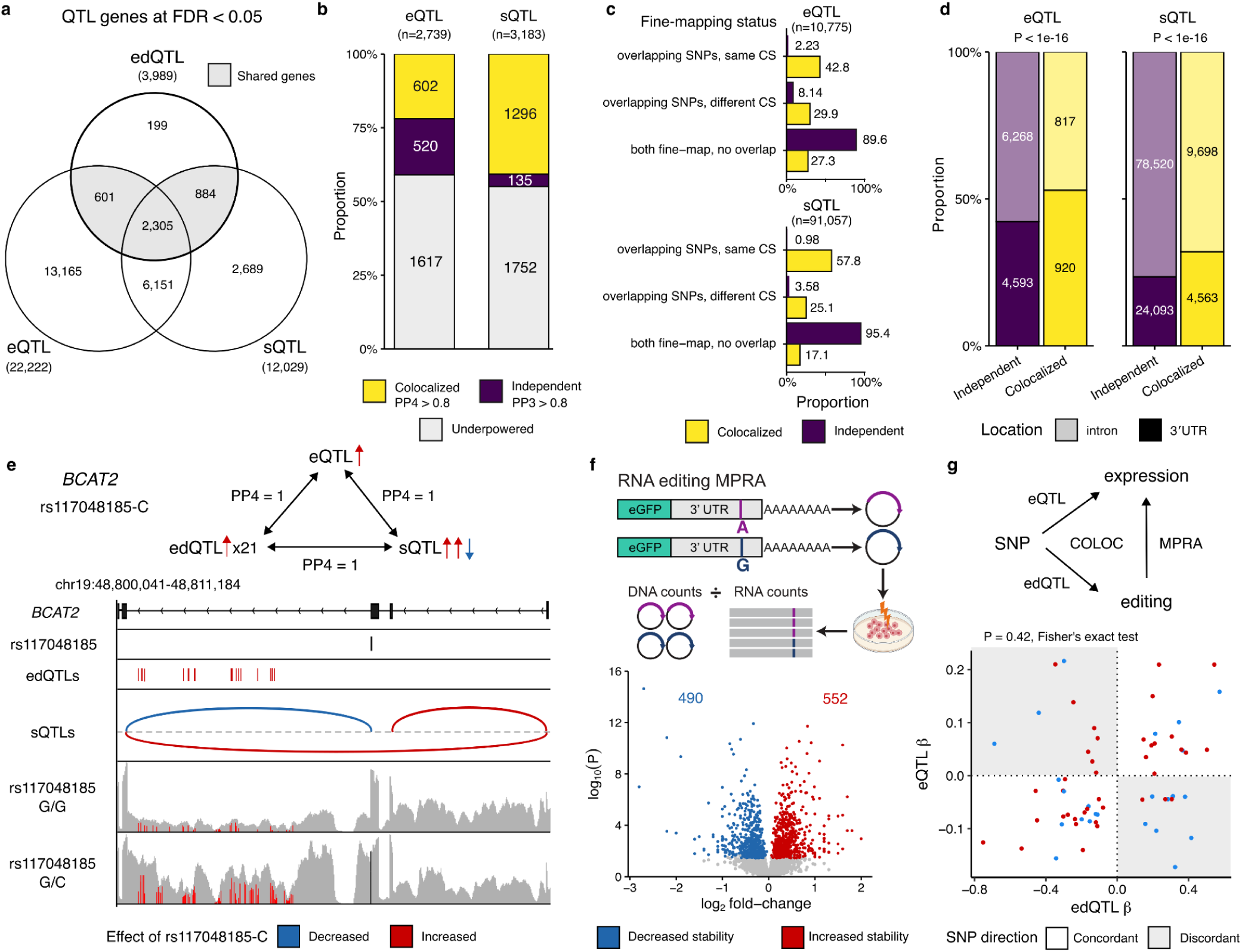
Sharing of genetic effects between edQTLs and other QTL types. **a)** All genes discovered (FDR 5%) between edQTLs, eQTLs, and sQTLs. edQTL gene assignment extended to include distal 3’UTRs. **b)** Genetic colocalization between edQTLs and eQTLs (left) and edQTLs and sQTLs (right) at the gene level. Highest PP4 reported for each gene across all tested edSite-gene and edSite-junction pairs. **c)** Colocalized edSites share fine-mapped SNPs with eQTLs and sQTLs in the same or different credible sets (CS) across all fine-mapped edSite-gene and edSite-junction pairs. **d)** Colocalized edSites are enriched within 3’UTRs. P < 1e-16, Fisher’s exact test for all edSite-gene and edSite-junction pairs. **e)** BCAT2 contains all three types of QTL associated with the same SNP. Upper panel shows relationships between rs117048185 and the three QTL types, including 21 edSites and 3 splice junction sQTLs. Lower panel shows genomic location of lead SNP, edSites and splice junctions. Representative IGV traces from a G/G homozygote and a G/C heterozygous donor, showing increased intron retention and corresponding A:I editing with the G/C allele. **f)** Meta-analysis significant edSites in 3’ UTRs were evaluated by MPRA to identify functional editing sites which alter RNA abundance. The volcano plot depicts expression fold-change (x-axis) and adjusted P-value (y-axis) on a logarithmic scale for each site tested, where colored dots denote sites that significantly (FDR 10%) altered mRNA abundance. **g)** Schematic of the relationship between QTLs, colocalization and MPRA. COLOC tests whether the same genetic variant (SNP) is associated with both editing and expression/splicing, whereas the MPRA tests whether the edited sequence alters mRNA stability. The sites with eQTL-edQTL colocalization tested in the MPRA are plotted to compare the QTL effects of the same SNP on editing rate (x-axis) and gene expression (y-axis), coloured by direction of stability change in the MPRA. PP: posterior probability.

For 168 genes all three QTL types colocalized to the same causal SNP (PP4 > 0.8 in all pairwise comparisons). One example is the mitochondrial gene branched-chain-amino-acid aminotransferase 2 (*BCAT2*), where all three QTL types share the lead SNP rs117048185, located within the third exon of the gene, and all three QTL types colocalized with each other with PP4 = 1. Strikingly, both edQTLs and sQTLs involve features within the first three introns of the gene. The minor C allele is associated with increased *BCAT2* expression, increased editing of 21 edSites within intron 3, and decreased usage of a splice junction that skips exons 2 and 3 (**Fig. 4e**). By comparing the RNA-seq read coverage over *BCAT2* in samples either homozygous for the reference G/G allele or heterozygous for the minor C allele, we observe that rs117048185-C carriers have increased retention of the intron 3 which contains the edSites. Therefore, it is likely that the change in RNA editing and gene expression captured by the edQTLs and eQTL respectively is driven by the rs117048185-C effect on intron 3 splicing, which acts to increase total *BCAT2* expression.

We performed an MPRA to test whether increased RNA editing can alter gene expression (**Fig. 4f**). Specifically, we tested the hypothesis that editing sites in 3’UTRs could influence gene expression by changing the sequence at locations where microRNAs (miRNAs) bind^29^. Using MapUTR^32^, we performed an *in vitro* screen for 3’UTR edSites that may alter gene expression through modulating miRNA binding. We performed *in silico* miRNA binding predictions using miRanda^33^, and 3,423 edSites within 3’UTRs were selected for testing if they were predicted to alter the sequence in the seed region of a predicted miRNA binding site, as this is the most critical region for miRNA target recognition. We performed an MPRA using the MapUTR platform^32^ to test whether these prioritized edSites destabilized expression of the 3’UTR sequence. The MPRA revealed 1,042 significantly changed sites (FDR 10%), representing edSites with potential functional relevance for gene expression regulation (**Fig. 4f**; **Table 5**). We then integrated our colocalized pairs of edQTLs and eQTLs, and compared the effect size (β) of the lead eQTL SNP on editing and expression with the direction of effect in the MPRA. We would expect genes with concordant β relationships (increased editing increases expression, or decreased editing decreases expression) to show increased stability in the MPRA (increased editing increases stability). However, we are underpowered to test this with only 61 gene-edSite pairs (P = 0.42, Fisher’s exact test), demonstrating the difficulty of testing this hypothesis.

### BigBrain edQTLs mediate neurodegenerative and psychiatric disorder risk

To evaluate the contributions of RNA editing to genetic risk for neurological disorders, we first tested whether our edQTLs explain disease heritability from GWAS using mediated expression score regression (MESC)^34^. While MESC showed no mediated edQTL heritability for height, we found evidence of non-zero mediation of heritability by edQTLs across neuropsychiatric disorders and neurodegenerative diseases, including schizophrenia^35,36^, bipolar disorder^37^, major depressive disorder (depression)^38^, multiple sclerosis (MS)^39^, amyotrophic lateral sclerosis (ALS)^40,41^, Alzheimer’s disease (AD)^42^, and Parkinson’s disease (PD)^43^ (**Fig. 5a**). Compared to psychiatric disorders, we observed elevated RNA editing mediated heritability for neurodegenerative diseases, highest in ALS (*h^2^_med_*/*h^2^_g_*= 0.21, **Fig. 5a**).

**Figure 5.**
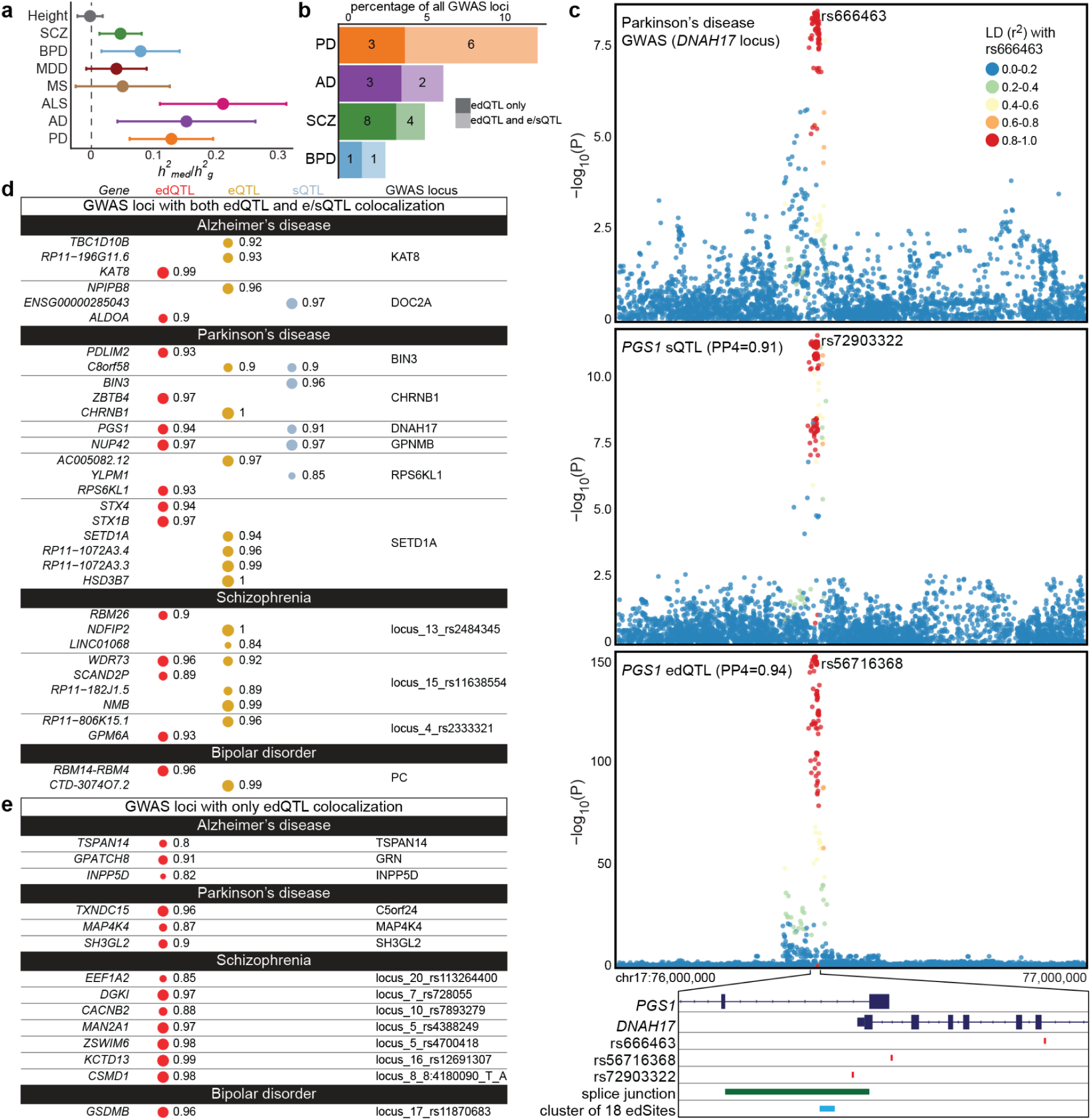
Contribution of edQTLs to genetic risk for neurological disorders. **a)** Estimated proportion of heritability mediated by edQTLs (h^2^_med_/h^2^_g_) for schizophrenia, bipolar disorder, major depressive disorder (MDD), multiple sclerosis (MS), amyotrophic lateral sclerosis (ALS), Alzheimer’s disease (AD), and Parkinson’s disease (PD). Height is included as a control. **b)** Proportion of all GWAS loci (x-axis) for a given neurological disease (y-axis) with strong evidence of colocalization with an edQTL. Colocalizations were only considered when the lead GWAS and QTL SNPs were within 500 Kb and had linkage disequilibrium (LD) r^2^ > 0.1. Strong evidence of colocalization was defined as: (i) PP4 > 0.8, and (ii) both GWAS and QTL SNPs were within 100 Kb of the editing site. The number of loci for each GWAS is provided to the right of each bar. **c)** LocusZoom (hg19) for an edQTL in the gene PGS1 (bottom panel) which colocalizes (PP4 = 0.94) with the DNAH17 PD locus (top panel). This GWAS locus also colocalizes with a splicing QTL (sQTL, middle panel) in PGS1 (PP4 = 0.91). Each dot, representing a SNP, is colored by LD with the GWAS index SNP (rs666463). Both the lead edQTL SNP (rs56716368) and lead sQTL SNP (rs72903322) are in perfect LD (r^2^ = 1) with rs666463. Below the locusZoom, an illustration of PGS1 and DNAH17 gene bodies (navy blue) highlights position of all edSites in PGS1 (light blue) as well as the sQTL splice junction (green) and the GWAS index as well as lead QTL SNPs (red). **d)** For all GWAS loci (right, loci nomenclature from each GWAS) with strong evidence of colocalization with an edQTL (red) if there was also a colocalizing eQTL (yellow) or sQTL (blue), all QTL genes (left, italicized) are reported with PP4. **e)** All GWAS loci (right, loci nomenclature from each GWAS) with only edQTL colocalization. PP: posterior probability.

Next, we performed colocalization analysis to test whether individual GWAS loci and edQTL share a causal variant^31,44^. We define strong evidence of colocalization when all three of the following criteria were met: (i) lead GWAS and edQTL SNPs are within 100 Kb or within 500 Kb and in moderate linkage disequilibrium (r^2^ > 0.1), (ii) both GWAS and edQTL SNPs are within 100 Kb of editing site, (iii) PP4 > 0.8. Across all traits tested, we identified a total of 75 edQTL with strong evidence of colocalization at 28 independent GWAS loci (**Fig. 5b**, **Table 6**). In three cases, the lead edQTL SNP was the GWAS index SNP, and in 24 cases the GWAS index SNP could be found in the same CS as the fine-mapped edQTL SNP.

The gene *PGS1* encodes phosphatidylglycerophosphate synthase 1, a mitochondrial inner-membrane enzyme involved in the synthesis of phosphatidylglycerol and cardiolipin. Cardiolipin is an essential phospholipid for mitochondrial function, dysregulation of which is a key component of PD etiology^45^. There is a cluster of 18 editing sites in the last intron of *PGS1*. edQTL signal for 10 of these sites colocalize with the *DNAH17* PD GWAS locus (**Fig. 5c**). Notably, these 18 edQTL also colocalize with a *PGS1* sQTL (PP4 = 0.93) and this sQTL also colocalizes with the GWAS locus (**Fig. 5c**). We identified 13 GWAS loci with strong evidence of edQTL colocalization where an eQTL or sQTL also colocalizes (**Fig. 5d**). At these loci, the edQTL almost always prioritizes a different causal gene than the e/sQTL, however there are three loci where both QTL genes are the same (**Fig. 5d**). There is strong evidence of edQTL and eQTL colocalization for the gene *WDR73* at the schizophrenia GWAS locus chr15:rs11638554 (**Fig. 5d**). Moreover, the *WDR73* eQTL and edQTL signals also colocalize (PP4 = 0.99). There is strong evidence of three-way colocalization of edQTL and sQTL for the gene *NUP42* and the PD GWAS locus *GPNMB* (**Fig. 5d**).

More than half (63%) of all GWAS loci with edQTL colocalization did not have e/sQTL colocalization signal, suggesting RNA editing mediates disease risk that cannot be explained by gene expression or splicing (**Fig. 5e**). A previous study using the CMC datasets found four schizophrenia loci colocalizing (PP4 > 0.5) with edQTL signal^7^. Here, we replicate the strongest colocalization signal from that study, of an editing site located slightly downstream of the 3’UTR of the gene *DGKI*, and identify an additional 11 schizophrenia loci with edQTL colocalization.

## Discussion

Regulation of A-to-I editing by cis-acting genetic variants (edQTL) has been explored in only a handful of studies to date, most notably with data from large consortia including GTEx^16,29^, CMC^7,46^, and AMP-AD^47^. In this study, part of the BigBrain Project, we present the largest RNA editing QTL mapping effort to date: a meta-analysis with a sample size (n = 10,202) orders of magnitude larger than previous studies, integrating all of these datasets with additional cohorts to reveal 38,731 edQTLs in 3,554 genes. We harmonized A-to-I editing events across 43 datasets, illuminating extensive heterogeneity and creating a valuable resource of high-confidence editing events. We leveraged colocalization analysis to evaluate the shared and distinct genetic regulation of gene expression, splicing, and RNA editing. Using an MPRA, we shortlisted putatively functional 3’UTR editing sites which may alter RNA abundance through interfering with miRNA targeting. Moreover, we described the portion of disease heritability for neurodegenerative diseases and psychiatric disorders mediated by edQTLs, and identified dozens of loci where RNA editing may act as the mechanism underlying disease risk. The scale and rigor of this work generated a resource which will serve as a platform for hypothesis generation and targeted validation studies to further our understanding of the biological underpinnings of brain disorders.

The BigBrain Project made use of publicly available data, which was not designed with the purpose of studying RNA editing in mind. Nevertheless, insights from this work are valuable for informing future works which aim to study RNA editing from RNA-sequencing data. We found polyA+ selected libraries yield fewer A-to-I sites than total RNA, presumably because the majority of ADAR-mediated editing takes place in the nucleus on pre-mRNA. It is known that the majority of A-to-I sites occur in introns, in line with the genome-wide distribution of *Alu* elements^48^, but in this study, we demonstrated the challenge this poses for between-study comparisons, as intronic sites are harder to reproduce because traditional RNA-seq is designed to capture mature mRNA. Techniques for studying nascent pre-mRNA could help resolve intronic A-to-I sites which may be consequential to other co- and post-transcriptional RNA processing events, such as alternative splicing. While detection of individual editing events is quite variable, it has been previously demonstrated that there is stronger replication at the gene-level, suggesting editing of individual transcripts is mechanistically more important than any individual site^49^. Accordingly, while we identified a significant edQTL for only 33% of all A-to-I sites tested, we found at least one edQTL for 69% of all genes with A-to-I sites.

There are two broad classifications of A-to-I editing, site-selective editing at an isolated adenosine and hyper-editing along stretches of neighboring adenosines on the same transcript^18,19^. Here, we focus only on selective editing and do not explicitly quantify clusters of hyper-editing, as this requires specific protocols for RNA-seq alignment^50^ and a main feature of the BigBrain Project is uniform data processing. Hyper-editing of dsRNAs by ADAR enzymes is promiscuous and the exact determinants of which adenosines are targeted are not clear^50–52^. Thus, *cis*-acting variants which alter double-strandedness might have seemingly indiscriminate effects on any individual editing site within a larger cluster, but when aggregated the direction and magnitude of that variant’s effect on editing within a gene may be more interpretable. In the example of edQTL colocalization in *PGS1*, 10 edSites all colocalize with the GWAS locus, perhaps representing the same effect.

Moreover, by the process of tandem alternative polyadenylation (APA), mRNAs from the same gene can have different length 3’UTR. Longer 3’UTRs are characteristic of the CNS, sometimes extending up to 10 Kb^22,23^. In the case where extended 3’UTR sequence overlaps a nearby inversely oriented gene, a long, perfect dsRNA structure could result which would be a highly preferential target for ADAR-mediated RNA editing^16^. At the *DNAH17* PD locus, some colocalizing edSites are called as T:C while others are called as A:G, potentially signaling the formation of a dsRNA substrate between the 3’UTR of *DNAH17* and *PGS1* intron.

Conventional bulk-tissue RNA-seq profiling confounds the discovery of transcriptomic signatures unique to distinct cell types. Still, the number and sample size of bulk-tissue RNA-seq datasets still considerably outnumbers that of single-cell RNA-seq. Moreover, calling RNA editing is particularly challenging in single-cell RNA-seq because of limitations such as reduced sequencing depth, 3’ coverage bias and smaller sample size^19^. Adopting the approach of a previous study, here we used fluorescence-activated nuclei sorting (FANS) derived cellular pools to assemble a catalog of cell-type specific editing sites which we used to annotate edSites from our meta-analysis^19^. From 38,731 edQTLs, we found 19% (n = 7,527) of the edSites were cell-type specific. One limitation of this strategy is editing sites which are highly specific to cell types which make up a very small proportion of bulk-brain tissue, such as microglia, which make up only 5-15%, are less likely to be present in the set of editing sites used in our meta-analysis^53^. Beyond the cellular specificity of the editing sites themselves, it is widely accepted that the regulatory effect of genetic variants can diverge between cell types^54^. Future work should consider edQTL mapping in sorted populations of disease-relevant cell types.

Discerning the functional relevance of RNA editing in non-coding regions of genes remains one of the greatest challenges for the field. Identifying edQTLs which are also e/sQTLs represents an opportunity to prioritize editing events which may be consequential for the expression or splicing of a gene. A previous edQTL study in GTEx reported only modest overlap between edQTLs and eQTLs and sQTLs, suggesting that most edQTLs represent independent genetic effects from splicing or gene expression^16^. Using colocalization analysis, we demonstrated shared genetic regulation of RNA editing and gene expression for 15% of genes containing edQTLs. Consistent with GTEx, we observed a higher overlap between RNA editing and splicing (32%). We identified 168 genes where the same genetic effect putatively regulates RNA editing, gene expression, and splicing, such as the example of *BCAT2* where editing and expression levels increase along with an intron retention event. However, *BCAT2* exemplifies the difficulty of assigning a causal ordering to these molecular events, as colocalization does not itself test for mediation (one QTL drives the other), nor distinguishes causality from pleiotropy (the same SNP independently causes both QTLs).

Here, we also evaluated the shared genetic effects between RNA editing and GWAS loci for neurological disorders by colocalization analysis. At GWAS loci where both edQTLs and e/sQTLs colocalize, we found they tend to prioritize different causal genes, and found only three instances of overlap at the gene-level: *WDR73* (ed/eQTL), *NUP42* (ed/sQTL), and *PGS1* (ed/sQTL). We identified 15 GWAS loci which colocalize solely with edQTL, suggesting that RNA editing is a mechanism that may resolve missing GWAS heritability not explained by expression or splicing QTLs.

A-to-I editing regulates innate immunity through interferon response pathways, and it has been shown that edQTLs explain a considerable fraction of genetic risk for inflammatory diseases^16,55,56^. Studies of AD, PD, and ALS suggest neuroinflammation is not purely a consequence of neurodegeneration, but may play a causal role in disease progression^57,58^. In this study, we found RNA editing mediated disease heritability is higher for neurodegenerative diseases, relative to psychiatric disorders. A similar relationship was highlighted across diseases in the proportion of all GWAS loci with edQTL colocalization. The role of RNA editing in the pathophysiology of neurodegenerative disorders warrants further investigation.

## Methods

Additional methods for analyses shared across BigBrain Project meta-analysis are provided in the **Supplementary Note**.

### Identification and quantification of RNA editing

RNA editing was called individually for samples from each tissue within every cohort. JACUSA^59^ v.2.0.1 was leveraged for *de novo* calling (call-1 -a D,M,Y,E -P UNSTRANDED -F 1024 -R -s -m 20) and supervised calling (call-1 -a D,Y -P UNSTRANDED -F 1024 -A -s -m 20) from a list of known A-to-I editing sites hosted in the REDIportal database^26^. *De novo* calling of editing sites did not overlap homopolymeric or ENCODE’s blacklisted regions of hg38^60^. The resulting sites were first filtered within individual samples based on the following criteria: 10 reads must cover each *de novo* called site, with a lower threshold of 5 reads for sites from supervised calling; 3 “edited” reads must support each editing event; multiallelic sites were removed. Following individual sample-level filtering, sites were concatenated to a cohort-level matrix and retained if they were present in at least 10 samples with a mean editing rate > 1%.

Using ANNOVAR^61^, editing sites were annotated with conservation metrics using the phastConsElements30way table of the UCSC Genome Browser^62^, assigned to genes and genic regions with RefGene (2019 release) (intergenic sites were assigned to the nearest gene within 10 Kb), and annotated to repetitive regions using RepeatMasker v.4.1.161. ANNOVAR was additionally used to cross-reference editing sites against dbSNP^63^ v.153 and gnomAD^64^ and were removed if they overlapped any rare or common genetic variants with a minor allele frequency (MAF) > 5%. Additionally, gene symbols were manually updated to GENCODE v46. Then, sites annotated as T:C by ANNOVAR were overlapped with 3’UTR sequences from GENCODE v46, extended by 10kb downstream. For these sites, both genes were listed.

This filtered set of editing sites is then used in a second round of supervised calling by Jacusa2, in order to resolve true missingness from low coverage of editing sites within a given sample. Editing sites were required to have non-NA coverage in > 50% of samples, and samples were required to have non-NA coverage of > 20% of all editing sites in the final matrix. Only A-to-I sites, represented in the RNA-seq data by A-to-G substitution, were considered in downstream analyses. T-to-C substitutions annotated to negative strand genes were also considered A-to-I events^21^.

### Annotation of cell-type specific RNA editing sites

For a subset of samples (n=41) from the MSBB dataset (syn2580853; see **Supplementary Note**), fluorescence activated nuclei sorting (FANS) was used to isolate three cell types from the parahippocampal gyrus (BA36)^25^. The neuronal marker NeuN was used to isolate neurons from non-neuronal cells, which were further sorted based on expression of the oligodendrocyte marker SOX10. The three resultant cellular populations were: neurons (NeuN^+^/SOX10^-^), oligodendrocytes (NeuN^-^/SOX10^+^), and pooled microglia/astrocytes (NeuN^-^/SOX10^-^).

RNA editing was called using the JACUSA2 pipeline described above. Stringent criteria was applied downstream on the called editing sites to define “cell-type specific” editing sites. A mean editing rate of 5% was required and the site had to be present in > 80% of samples within their respective FANS-derived cellular pools, which will hereby be referred to as NEU (neurons), OLIG (oligodendrocytes), and MGAS (microglia/astrocytes). Furthermore, only sites which passed these criteria in one cellular pool and were not present in the other two were defined as cell-type specific in MSBB.

At this point, the list of NEU, OLIG, and MGAS cell-specific editing sites was merged with a previously published list of cell-specific editing sites from FANS-derived pools of excitatory (glutamatergic, or GLU) neurons, inhibitory (GABAergic, or GABA) neurons, and oligodendrocytes (OLIG) sorted from the prefrontal cortex^19^. The resultant combined list consists of six categorizations of cell-type annotations for editing sites: 1) “OLIG” sites had to be present in the oligodendrocyte pools from both experiments; 2) “MGAS” sites were only present in the MSBB list; 3) “GLU” sites were specific to the GLU pool in *Cuddleston et al.* and also replicate in the NEU pool from MSBB; 4) “GABA” sites were specific to the GABA pool in *Cuddleston et al.* and replicate in the NEU pool from MSBB; 5) “GABA-GLU” were sites enriched in both the GABA and GLU pools in *Cuddleston et al.* and replicate in the NEU pool from MSBB; 6) “NEU” sites passed filtering criteria to be defined as cell-specific in MSBB but did not replicate in the neuronal pools from *Cuddleston et al*.

### Mapping RNA editing QTLs

An editing ratio, defined as the number of edited reads divided by the total number of reads covering an A-to-I site, was used as the phenotype for QTL mapping. Editing sites with no variance between samples were removed. Moreover, to reduce the testing space and prioritize moderate effects, only editing sites with a mean editing rate > 5% present in > 75% of samples in each study were retained. Missing values in each tissue-cohort pair phenotype matrix were imputed with the median editing rate. Phenotype matrices were harmonized across datasets, including any editing site present in 2 or more studies. The phenotype matrix was normalized (see **Supplementary Note**) and known (age, biological sex, and VOOM normalized *ADAR*/*ADARB1* expression) and hidden (top 5 PEER factors^65^) covariates were included for QTL mapping and meta-analysis with mmQTL^17^ (see **Supplementary Note**).

### *In silico* miRNA binding predictions

For each 3’UTR editing site with a meta-analysis significant edQTL association, genomic sequences for +/- 10 bp surrounding the editing site were obtained using BEDTools^66^ getFasta. Both an edited and unedited version of the 11 bp sequences was created. Local alignments for these sequences were tested against all mature miRNAs in miRbase^67^ using the tool miRANDA^33^ (using the -strict flag), which implements a position-weighted alignment algorithm and reports a score of sequence complementarity along with the stability of the resulting RNA duplex and minimum free energy (MFE). Only miRNA-UTR hits with a score > 145 and MFE < -20 were considered.

### 3’UTR massively parallel reporter assay

3’UTR edSites prioritized by *in silico* miRNA binding predictions were tested to determine whether they had an effect on mRNA abundance *in vitro* using a massively parallel reporter assay (MPRA) approach with MapUTR^32^. The design uses a library of 164 bp oligomers cloned into a reporter vector with either the edited or unedited sequence. Reporter plasmids carrying the same 3’UTR sequence, differing only at the editing site, were electroporated into HEK293T cells. 24 hours later, the mRNA was extracted and RNA-sequencing libraries were prepared for three biological replicates. In parallel, DNA-sequencing libraries were also prepared from the plasmid library to be used for RNA normalization. After sequencing, MPRAnalyze^68^ was used to identify sites with a differential RNA/DNA ratio in the edited sequence relative to the unedited. Significance was considered at a 10% false discovery rate.

### QTL-QTL Colocalization

All pairs of QTL features (editing sites, genes, and junctions) passing significance testing were matched for pairwise comparisons using COLOC^31^ (v5.2.3). COLOC estimates 5 posterior probabilities, including probability of both QTLs being independent (PP3) and colocalized (PP4). The threshold for colocalization was set at PP4 > 0.8 and independence at PP3 > 0.8.

### Statistics & Reproducibility

No statistical method was used to predetermine sample size. No data were excluded from the analyses. The experiments were not randomized. The investigators were not blinded to allocation during experiments and outcome assessment. For all QTL mapping, data distribution was assumed to be normal after log-normal transformation, but this was not formally tested.

## Data Availability

No sequencing data was generated as part of this project. All summary statistics from QTL analyses are publicly available on Zenodo (10.5281/zenodo.17210092). Data accession numbers for each cohort are listed in the Supplementary Note. We provide an interactive browser for all BigBrain QTLs at https://bigbrain.nygenome.org

https://zenodo.org/records/17210092

https://zenodo.org/records/17153730

https://zenodo.org/records/17226890

https://zenodo.org/records/17297440

## Acknowledgements

We thank the patients and families for their generous gift of brain donation. We also thank the investigators and consortia who generated and shared the RNA-seq datasets analyzed here. We have acknowledged each of the included cohorts and datasets in the **Supplementary Note**.

This study was supported by the following National Institutes of Health grants: NIA U01-AG068880, NIA R21-AG063130, NIA R01-AG054005, NIA RF1-AG065926, NIA R01-AG065926 NIA R56-AG088669, NIA R21-AG091272, NIA P30-AG066514, NINDS U54-NS123743, and NINDS R01-NS116006 to TR, JH, BZM, KBP, BJ, and WHC; NIA U01-AG068880 to TR, JH, BZM, KBP, BJ, WHC, AR, and DAK; NIA F31-AG084223 to WHC; NIH R01-MH123177 to XX.

This work was supported in part through the computational resources and staff expertise provided by Scientific Computing at the Icahn School of Medicine at Mount Sinai and supported by the Clinical and Translational Science Awards (CTSA) grant UL1TR004419 from the National Center for Advancing Translational Sciences. Research reported in this paper was supported by the Office of Research Infrastructure of the National Institutes of Health under award number S10OD026880 and S10OD030463. The content is solely the responsibility of the authors and does not necessarily represent the official views of the National Institutes of Health.

## Data availability

No sequencing data was generated as part of this project. All summary statistics from QTL analyses are publicly available on Zenodo (10.5281/zenodo.17210092). Data accession numbers for each cohort are listed in the **Supplementary Note**. We provide an interactive browser for all BigBrain QTLs at https://bigbrain.nygenome.org

## Code Availability

Genotype QC: https://github.com/RajLabMSSM/Genotype_QC_Pipeline_2.0

RNA editing: https://github.com/RajLabMSSM/editing-pipeline/tree/master/jacusa-pipeline

mmQTL: https://github.com/RajLabMSSM/mmQTL-pipeline

MESC: https://github.com/RajLabMSSM/downstream-QTL/tree/master/MESC

COLOC: https://github.com/RajLabMSSM/downstream-QTL/tree/master/COLOC

## Author Contributions

The study was conceived by JH and TR. Data analysis was led by WHD, with contributions from JH, KBP, BZM, BJ, HS, AR, MSB, and DAK. Data generation was performed by TF and XGX. Work was supervised by JH, DAK, HHW, XGX, and TR. The manuscript was written by WHD and revised by JH and TR, with input from all co-authors.

---

**Supplementary Figure 1.**
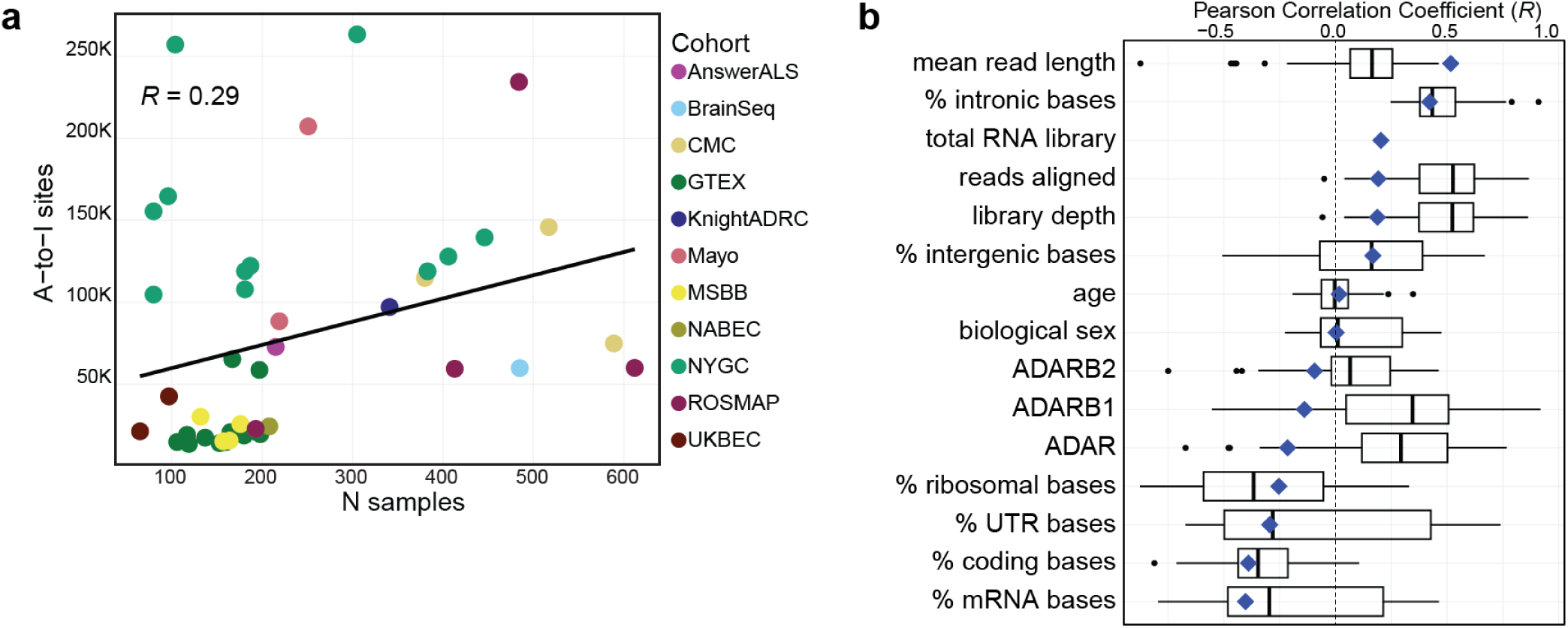
Variance in editing site discovery within and between 43 datasets. **a)** Pearson correlation (*R*) between the number of A-to-I sites (y-axis) which passed filtering thresholds in each dataset (tissue-cohort pair) by dataset sample size (x-axis). Points are colored by the cohort each dataset belongs to. **b)** Boxplots represent the distribution of Pearson correlation coefficients (x-axis) from associations run within each dataset evaluating the correlation between each variable (y-axis) and the number of editing sites which passed filtering thresholds in individual samples in that dataset. Boxplots plot the first quartile, the median and the third quartile of the values, with the whiskers denoting 1.5 times the interquartile range. Black dots represent outliers defined by 1.5 times the interquartile range. Vertical dashed black line denotes *R*=0. The blue diamond overlapping each respective boxplot represents the correlation coefficient (*R*) for the variable against the mean discovery rate in each dataset.

**Supplementary Figure 2.**
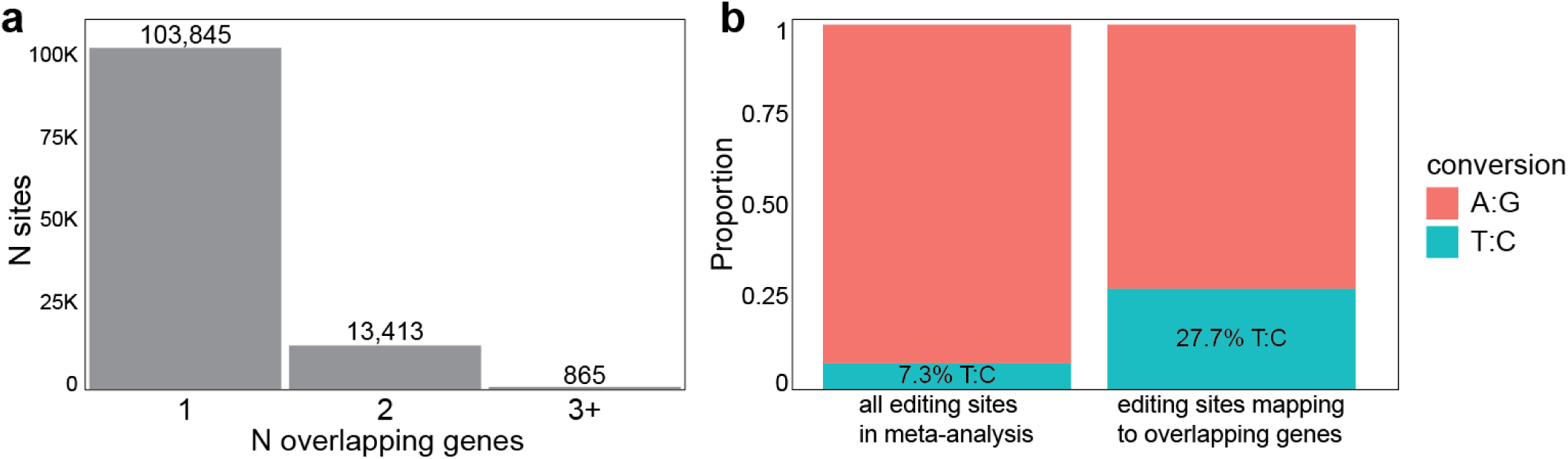
Antisense A-to-I editing. **a)** The number of editing sites which overlap multiple GENCODE v46 transcripts. **b)** Proportion of T:C conversions amongst all editing sites included in the meta-analysis versus those sites which overlap inversely oriented genes.

**Supplementary Figure 3.**
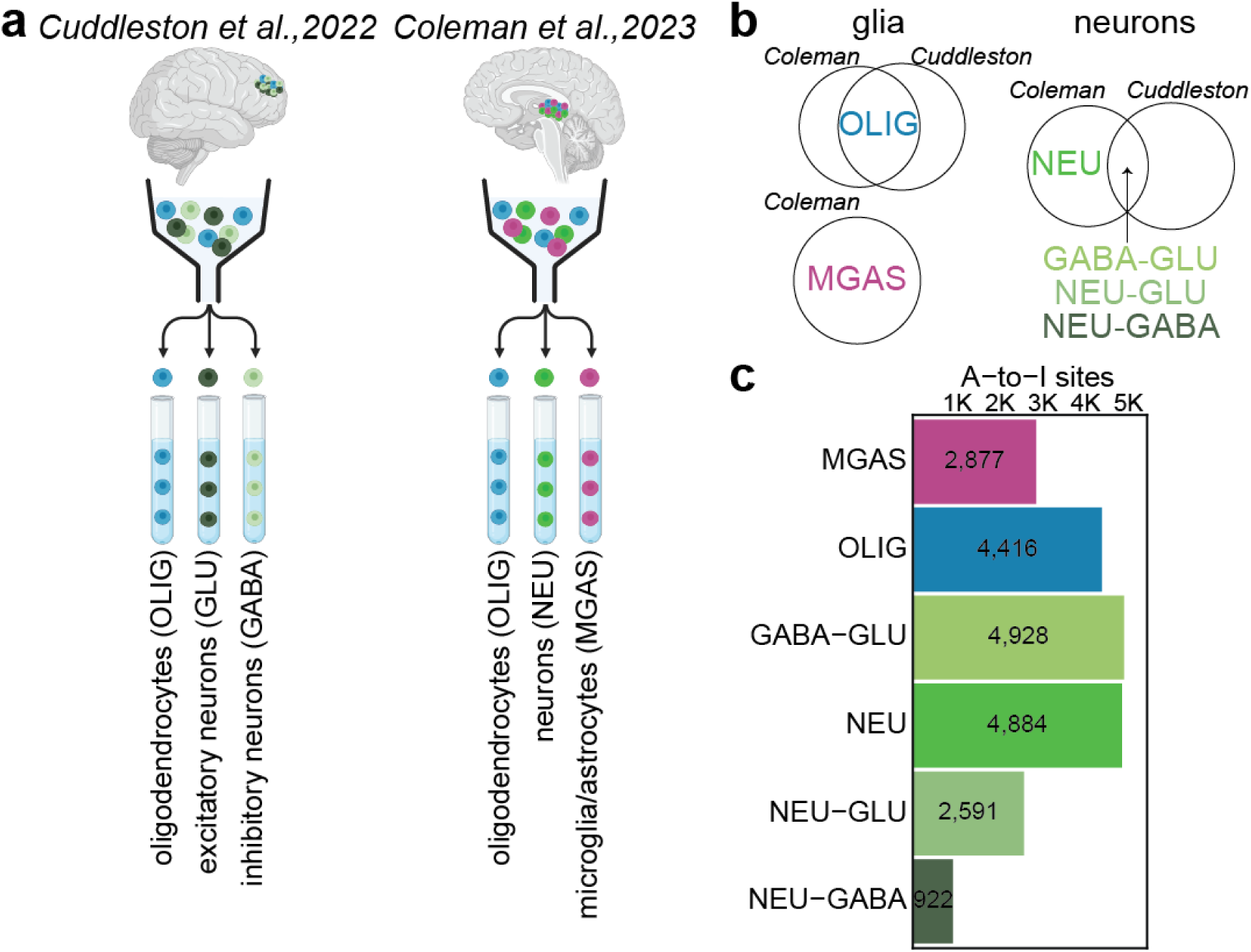
Cellular specificity of A-to-I sites in BigBrain. **a)** Depiction of the fluorescence-activated nuclei sorting (FANS) strategies from *Cuddleston et al., 2022* (*Cuddleston*) and *Coleman et al., 2023* (*Coleman*). In *Cuddleston*, three pools were sorted from prefrontal cortex tissue: oligodendrocytes (NeuN^-^/SOX10^+^), excitatory GABAergic neurons (NeuN^+^/SOX6^+^), and inhibitory glutamatergic neurons (NeuN^+^/SOX6^-^). In *Coleman*, three pools were sorted from parahippocampal gyrus tissue: oligodendrocytes (NeuN^-^/SOX10^+^), neurons (NeuN^+^/SOX10^-^), and pooled microglia/astrocytes (NeuN^+^/SOX10^+^). **b)** Editing sites were defined as cell-type specific within each study, and these categories were harmonized for cellular annotation of the sites identified in BigBrain (see Methods), where MGAS were distinct in *Coleman*; OLIG replicated in both studies; NEU were distinct in *Coleman*; GABA-GLU were sites shared by the GABA and GLU pools in *Cuddleston* which replicate in the NEU pool in *Coleman*; NEU-GLU and NEU-GABA were specific to their respective pools in *Cuddleston* and replicate in the NEU pool in *Coleman*. **c)** Of the 118,123 high-confidence A-to-I sites, the number of sites (x-axis) annotated to each cell type (y-axis) are represented by colored bars. MGAS: microglia/astrocytes; OLIG: oligodendrocytes; NEU: neurons; GLU: glutamatergic neurons; GABA: GABAergic neurons.

**Supplementary Figure 4.**
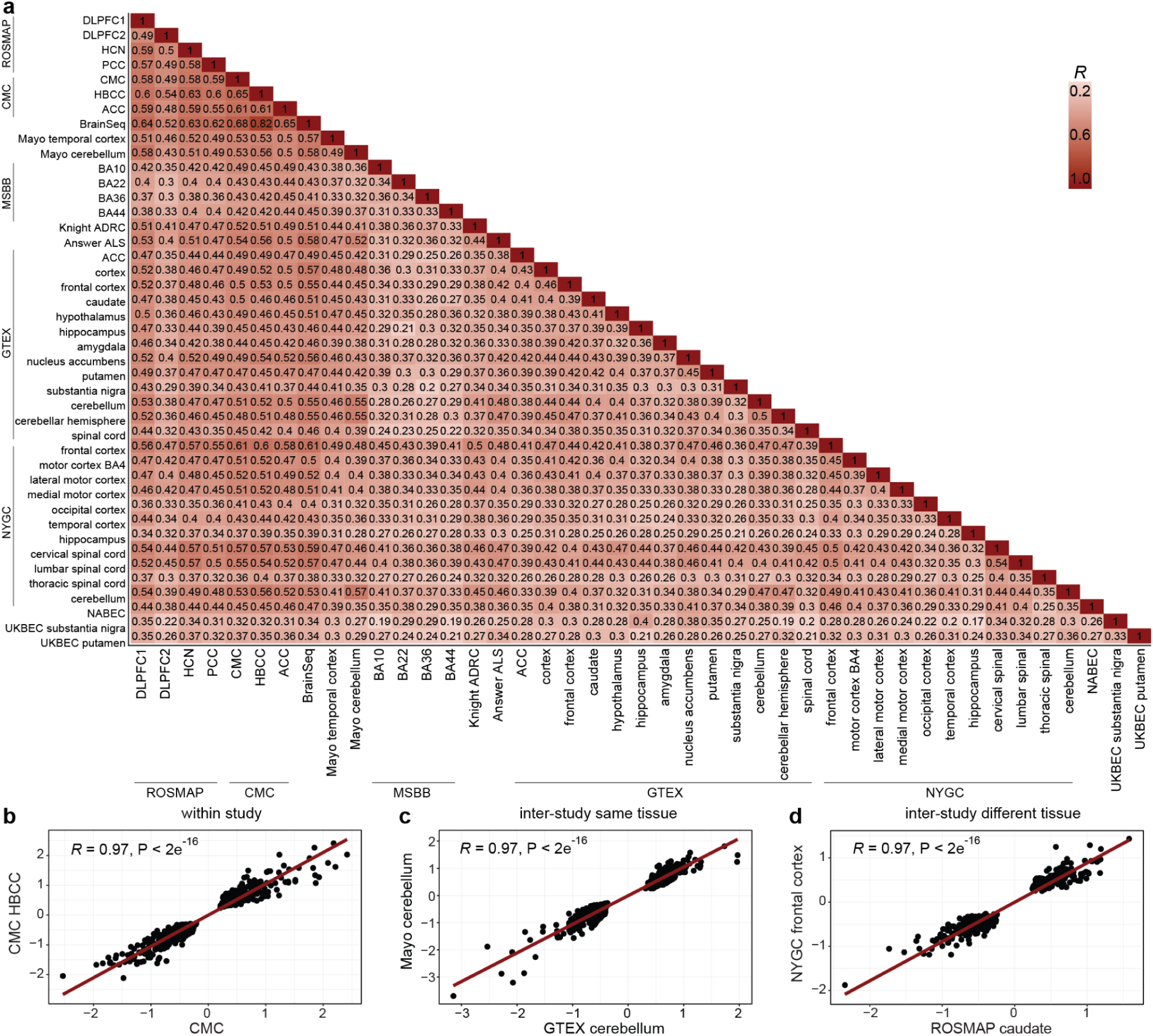
Correlation of edQTL effect sizes between BigBrain datasets. **a)** Pairwise correlation (Pearson) tests for all BigBrain datasets which were meta-analyzed. The heatmap is colored by the estimate (*R*), which is also displayed in each tile. Scatterplots highlight select correlations between two datasets which were from **b)** the same cohort, **c)** the same tissue across different cohorts, and **d)** different tissues from different cohorts, where the red trendline corresponds to the estimate and p-values displayed on each plot from a two-sided Pearson correlation test.

**Supplementary Figure 5.**
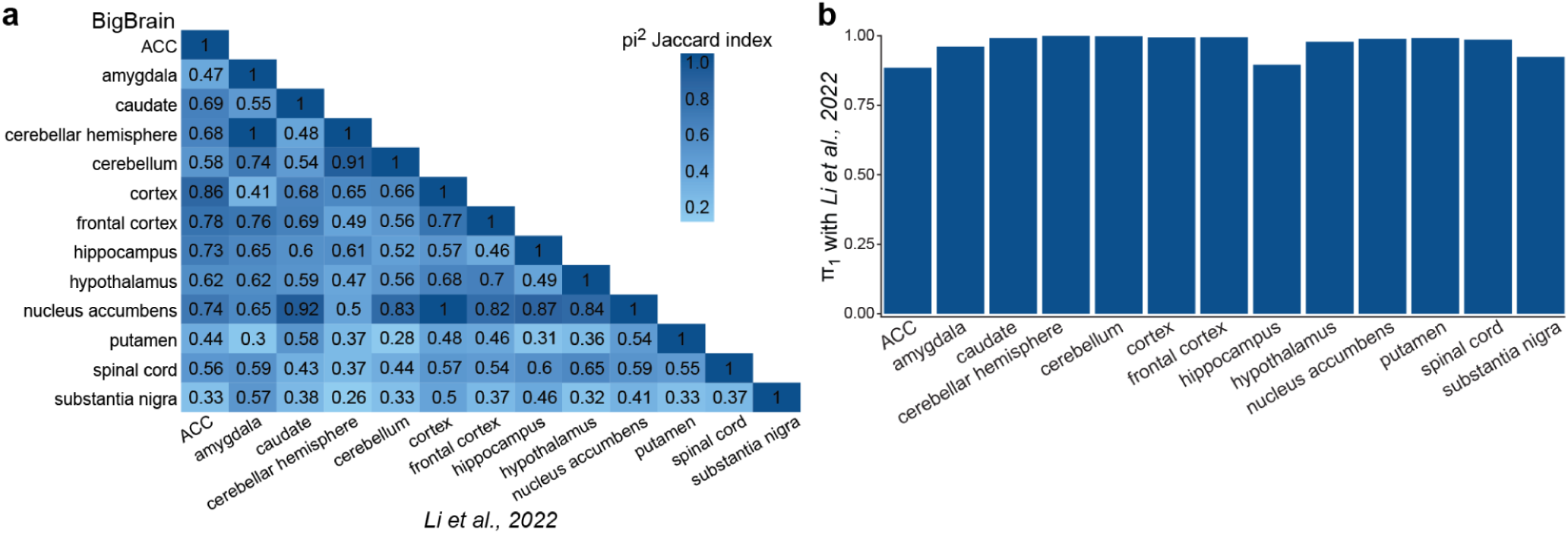
Replication of previously published edQTLs in BigBrain. **a)** π^2^ analysis between all GTEx tissues from BigBrain (y-axis) and *Li et al., 2022* (x-axis). The heatmap is colored by Jaccard similarity index. **b)** Storey’s π_1_ replication (y-axis) of *Li et al., 2022* edQTLs in BigBrain for each GTEx tissue (x-axis).

**Supplementary Figure 6.**
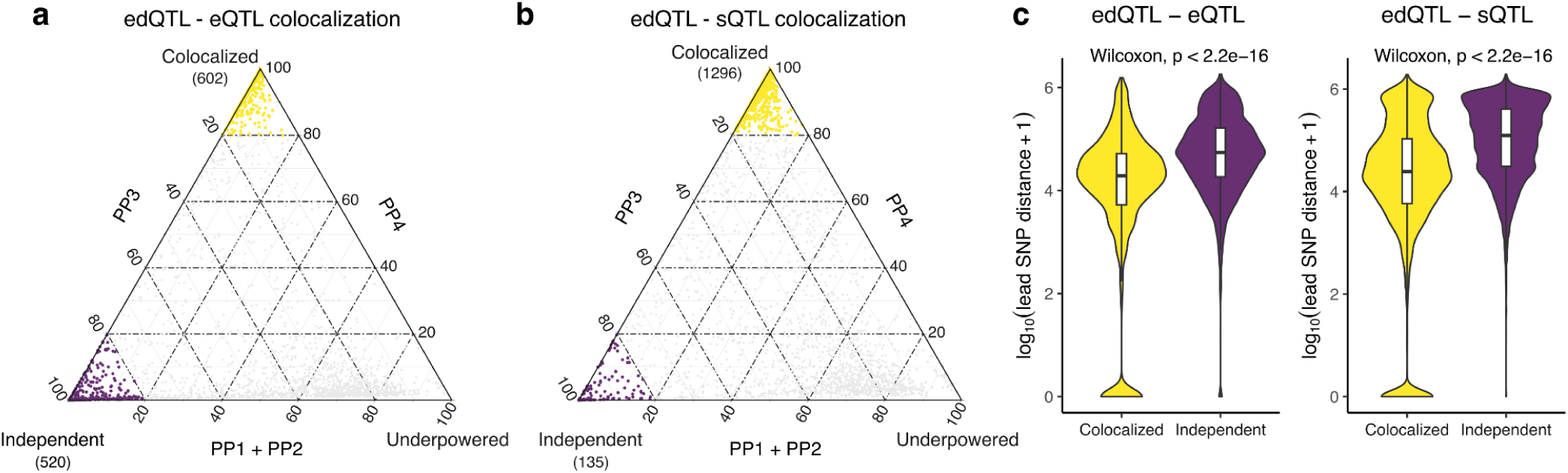
Sharing of QTLs across editing, expression and splicing. Ternary plots showing the posterior probabilities for pairs of **a)** edQTLs and eQTLs and **b)** edQTLs with sQTLs. PP3 is the posterior probability of the two QTL types being independent, and PP4 is the posterior probability of the two QTL types sharing the same single variant. **c)** Distance between the lead SNPs for pairs of QTLs.

## References

1. Maurano, M. T. et al. Systematic localization of common disease-associated variation in regulatory DNA. Science 337, 1190–1195 (2012).

2. Aguet, F. et al. The GTEx Consortium atlas of genetic regulatory effects across human tissues. SCIENCE 369, 1318–1330 (2020).

3. Nicolae, D. L. et al. Trait-associated SNPs are more likely to be eQTLs: annotation to enhance discovery from GWAS. PLoS Genet. 6, e1000888 (2010).

4. Mostafavi, H., Spence, J. P., Naqvi, S. & Pritchard, J. K. Systematic differences in discovery of genetic effects on gene expression and complex traits. Nat. Genet. 55, 1866–1875 (2023).

5. Cappannini, A. et al. MODOMICS: a database of RNA modifications and related information. 2023 update. Nucleic Acids Res. 52, D239–D244 (2024).

6. Behm, M. & Öhman, M. RNA editing: A contributor to neuronal dynamics in the mammalian brain. Trends Genet. 32, 165–175 (2016).

7. Breen, M. S. et al. Global landscape and genetic regulation of RNA editing in cortical samples from individuals with schizophrenia. Nat. Neurosci. 22, 1402–1412 (2019).

8. Choudhury, M. et al. Widespread RNA hypoediting in schizophrenia and its relevance to mitochondrial function. Sci. Adv. 9, eade9997 (2023).

9. Eran, A. et al. Comparative RNA editing in autistic and neurotypical cerebella. Mol. Psychiatry 18, 1041–1048 (2013).

10. Tran, S. S. et al. Widespread RNA editing dysregulation in brains from autistic individuals. Nat. Neurosci. 22, 25–36 (2019).

11. Ma, Y. et al. Atlas of RNA editing events affecting protein expression in aged and Alzheimer’s disease human brain tissue. Nat. Commun. 12, 7035 (2021).

12. Niswender, C. M., Copeland, S. C., Herrick-Davis, K., Emeson, R. B. & Sanders-Bush, E. RNA editing of the human serotonin 5-hydroxytryptamine 2C receptor silences constitutive activity. J. Biol. Chem. 274, 9472–9478 (1999).

13. Hoopengardner, B., Bhalla, T., Staber, C. & Reenan, R. Nervous system targets of RNA editing identified by comparative genomics. Science 301, 832–836 (2003).

14. Bhalla, T., Rosenthal, J. J. C., Holmgren, M. & Reenan, R. Control of human potassium channel inactivation by editing of a small mRNA hairpin. Nat. Struct. Mol. Biol. 11, 950–956 (2004).

15. Bazak, L. et al. A-to-I RNA editing occurs at over a hundred million genomic sites, located in a majority of human genes. Genome Res. 24, 365–376 (2014).

16. Li, Q. et al. RNA editing underlies genetic risk of common inflammatory diseases. Nature 608, 569–577 (2022).

17. Zeng, B. A. et al. Multi-ancestry eQTL meta-analysis of human brain identifies candidate causal variants for brain-related traits. Nat. Genet. 54, 161–+ (2022).

18. Cuddleston, W. H. et al. Spatiotemporal and genetic regulation of A-to-I editing throughout human brain development. Cell Rep. 41, 111585 (2022).

19. Cuddleston, W. H. et al. Cellular and genetic drivers of RNA editing variation in the human brain. Nat. Commun. 13, 2997 (2022).

20. Gabay, O. et al. Landscape of adenosine-to-inosine RNA recoding across human tissues. Nat. Commun. 13, 1184 (2022).

21. Pecori, R. et al. ADAR RNA editing on antisense RNAs results in apparent U-to-C base changes on overlapping sense transcripts. Front. Cell Dev. Biol. 10, 1080626 (2022).

22. Bae, B. & Miura, P. Emerging roles for 3’ UTRs in neurons. Int. J. Mol. Sci. 21, 3413 (2020).

23. Miura, P., Shenker, S., Andreu-Agullo, C., Westholm, J. O. & Lai, E. C. Widespread and extensive lengthening of 3’ UTRs in the mammalian brain. Genome Res. 23, 812–825 (2013).

24. Wang, K., Li, M. & Hakonarson, H. ANNOVAR: Functional annotation of genetic variants from next-generation sequencing data. Nucleic Acids Research 38, (2010).

25. Coleman, C. et al. Multi-omic atlas of the parahippocampal gyrus in Alzheimer’s disease. Sci. Data 10, 602 (2023).

26. Picardi, E., D’Erchia, A. M., Lo Giudice, C. & Pesole, G. REDIportal: a comprehensive database of A-to-I RNA editing events in humans. Nucleic Acids Res. 45, D750–D757 (2017).

27. Storey, J. D. & Tibshirani, R. Statistical significance for genomewide studies. Proc. Natl. Acad. Sci. U. S. A. 100, 9440–9445 (2003).

28. Knowles, D. A. et al. Determining the genetic basis of anthracycline-cardiotoxicity by molecular response QTL mapping in induced cardiomyocytes. Elife 7, (2018).

29. Park, E., Jiang, Y., Hao, L., Hui, J. & Xing, Y. Genetic variation and microRNA targeting of A-to-I RNA editing fine tune human tissue transcriptomes. Genome Biol. 22, 77 (2021).

30. Réal, A. et al. Mapping genetic effects on splicing in ten thousand post-mortem brain samples reveals novel mediators of neurological disease risk. medRxiv (2025) doi:10.1101/2025.09.25.25336663.

31. Wallace, C. Statistical Testing of Shared Genetic Control for Potentially Related Traits. Genet. Epidemiol. 37, 802–813 (2013).

32. Fu, T. et al. Massively parallel screen uncovers many rare 3’ UTR variants regulating mRNA abundance of cancer driver genes. Nat. Commun. 15, 3335 (2024).

33. Enright, A. J. et al. MicroRNA targets in Drosophila. Genome Biol. 5, R1 (2003).

34. Yao, D. W., O’Connor, L. J., Price, A. L. & Gusev, A. Quantifying genetic effects on disease mediated by assayed gene expression levels. Nat. Genet. 52, 626–633 (2020).

35. Schizophrenia Working Group of the Psychiatric Genomics Consortium. Biological insights from 108 schizophrenia-associated genetic loci. Nature 511, 421–427 (2014).

36. Trubetskoy, V. et al. Mapping genomic loci implicates genes and synaptic biology in schizophrenia. Nature 604, 502–508 (2022).

37. Stahl, E. A. et al. Genome-wide association study identifies 30 loci associated with bipolar disorder. Nat. Genet. 51, 793–803 (2019).

38. Wray, N. R. et al. Genome-wide association analyses identify 44 risk variants and refine the genetic architecture of major depression. Nat. Genet. 50, 668–681 (2018).

39. International Multiple Sclerosis Genetics Consortium. Multiple sclerosis genomic map implicates peripheral immune cells and microglia in susceptibility. Science 365, eaav7188 (2019).

40. Nicolas, A. et al. Genome-wide analyses identify KIF5A as a novel ALS gene. Neuron 97, 1268–1283.e6 (2018).

41. van Rheenen, W. et al. Author Correction: Common and rare variant association analyses in amyotrophic lateral sclerosis identify 15 risk loci with distinct genetic architectures and neuron-specific biology. Nat. Genet. 54, 361 (2022).

42. Bellenguez, C. et al. New insights into the genetic etiology of Alzheimer’s disease and related dementias. Nat. Genet. 54, 412–436 (2022).

43. Nalls, M. A. et al. Identification of novel risk loci, causal insights, and heritable risk for Parkinson’s disease: a meta-analysis of genome-wide association studies. Lancet Neurol. 18, 1091–1102 (2019).

44. Giambartolomei, C. et al. Bayesian test for colocalisation between pairs of genetic association studies using summary statistics. PLoS Genet. 10, e1004383 (2014).

45. Paradies, G., Paradies, V., Ruggiero, F. M. & Petrosillo, G. Role of cardiolipin in mitochondrial function and dynamics in health and disease: Molecular and pharmacological aspects. Cells 8, 728 (2019).

46. Choudhury, M., Yamamoto, R. & Xiao, X. Genetic architecture of RNA editing, splicing and gene expression in schizophrenia. Hum. Mol. Genet. 34, 277–290 (2025).

47. Gupta, A. K. et al. Comprehensive characterization of the RNA editing landscape in the human aging brains with Alzheimer’s disease. Alzheimers. Dement. 21, e70452 (2025).

48. Grover, D., Mukerji, M., Bhatnagar, P., Kannan, K. & Brahmachari, S. K. Alu repeat analysis in the complete human genome: trends and variations with respect to genomic composition. Bioinformatics 20, 813–817 (2004).

49. Park, E., Williams, B., Wold, B. J. & Mortazavi, A. RNA editing in the human ENCODE RNA-seq data. Genome Res. 22, 1626–1633 (2012).

50. Porath, H. T., Carmi, S. & Levanon, E. Y. A genome-wide map of hyper-edited RNA reveals numerous new sites. Nat. Commun. 5, 4726 (2014).

51. Walkley, C. R. & Li, J. B. Rewriting the transcriptome: adenosine-to-inosine RNA editing by ADARs. Genome Biol. 18, 205 (2017).

52. Wahlstedt, H. & Ohman, M. Site-selective versus promiscuous A-to-I editing. Wiley Interdiscip. Rev. RNA 2, 761–771 (2011).

53. Thion, M. S., Ginhoux, F. & Garel, S. Microglia and early brain development: An intimate journey. Science 362, 185–189 (2018).

54. Cano-Gamez, E. & Trynka, G. From GWAS to function: Using functional genomics to identify the mechanisms underlying complex diseases. Front. Genet. 11, 424 (2020).

55. Mannion, N. M. et al. The RNA-editing enzyme ADAR1 controls innate immune responses to RNA. Cell Rep. 9, 1482–1494 (2014).

56. Liddicoat, B. J. et al. RNA editing by ADAR1 prevents MDA5 sensing of endogenous dsRNA as nonself. Science 349, 1115–1120 (2015).

57. Mayne, K., White, J. A., McMurran, C. E., Rivera, F. J. & de la Fuente, A. G. Aging and neurodegenerative disease: Is the adaptive immune system a friend or foe? Front. Aging Neurosci. 12, 572090 (2020).

58. Hammond, T. R., Marsh, S. E. & Stevens, B. Immune signaling in neurodegeneration. Immunity 50, 955–974 (2019).

59. Piechotta, M., Naarmann-de Vries, I. S., Wang, Q., Altmüller, J. & Dieterich, C. RNA modification mapping with JACUSA2. Genome Biol. 23, 115 (2022).

60. Amemiya, H. M., Kundaje, A. & Boyle, A. P. The ENCODE blacklist: Identification of problematic regions of the genome. Sci. Rep. 9, 9354 (2019).

61. Wang, K., Li, M. & Hakonarson, H. ANNOVAR: functional annotation of genetic variants from high-throughput sequencing data. Nucleic Acids Res. 38, e164 (2010).

62. Karolchik, D. et al. The UCSC Table Browser data retrieval tool. Nucleic Acids Res. 32, D493–6 (2004).

63. Sherry, S. T. et al. dbSNP: the NCBI database of genetic variation. Nucleic Acids Res. 29, 308–311 (2001).

64. Karczewski, K. J. et al. Author Correction: The mutational constraint spectrum quantified from variation in 141,456 humans. Nature 590, E53 (2021).

65. Stegle, O., Parts, L., Piipari, M., Winn, J. & Durbin, R. Using probabilistic estimation of expression residuals (PEER) to obtain increased power and interpretability of gene expression analyses. Nat. Protoc. 7, 500–507 (2012).

66. Quinlan, A. R. & Hall, I. M. BEDTools: a flexible suite of utilities for comparing genomic features. Bioinformatics 26, 841–842 (2010).

67. Kozomara, A., Birgaoanu, M. & Griffiths-Jones, S. miRBase: from microRNA sequences to function. Nucleic Acids Res. 47, D155–D162 (2019).

68. Ashuach, T. et al. MPRAnalyze: statistical framework for massively parallel reporter assays. Genome Biol. 20, (2019).

