## Supplementary Figures for "Meta-analysis of genetic regulation of RNA editing in the human brain identifies new genes underlying neurological disease"

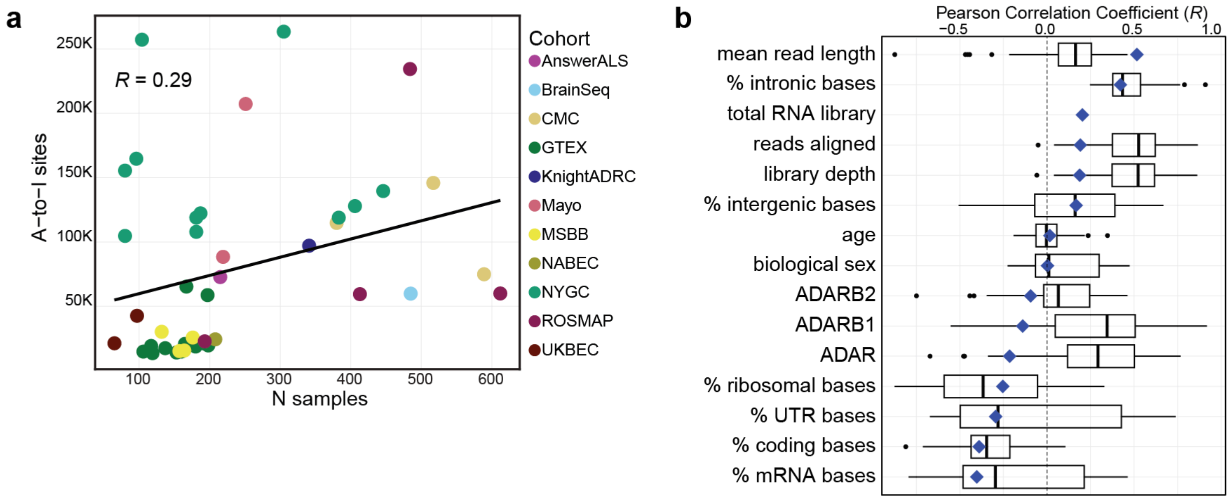


**Supplementary Figure 1 | Variance in editing site discovery within and between 43 datasets. a)** Pearson correlation (*R*) between the number of A-to-I sites (y-axis) which passed filtering thresholds in each dataset (tissue-cohort pair) by dataset sample size (x-axis). Points are colored by the cohort each dataset belongs to. **b)** Boxplots represent the distribution of Pearson correlation coefficients (x-axis) from associations run within each dataset evaluating the correlation between each variable (y-axis) and the number of editing sites which passed filtering thresholds in individual samples in that dataset. Boxplots plot the first quartile, the median and the third quartile of the values, with the whiskers denoting 1.5 times the interquartile range. Black dots represent outliers defined by 1.5 times the interquartile range. Vertical dashed black line denotes *R*=0. The blue diamond overlapping each respective boxplot represents the correlation coefficient (*R*) for the variable against the mean discovery rate in each dataset.

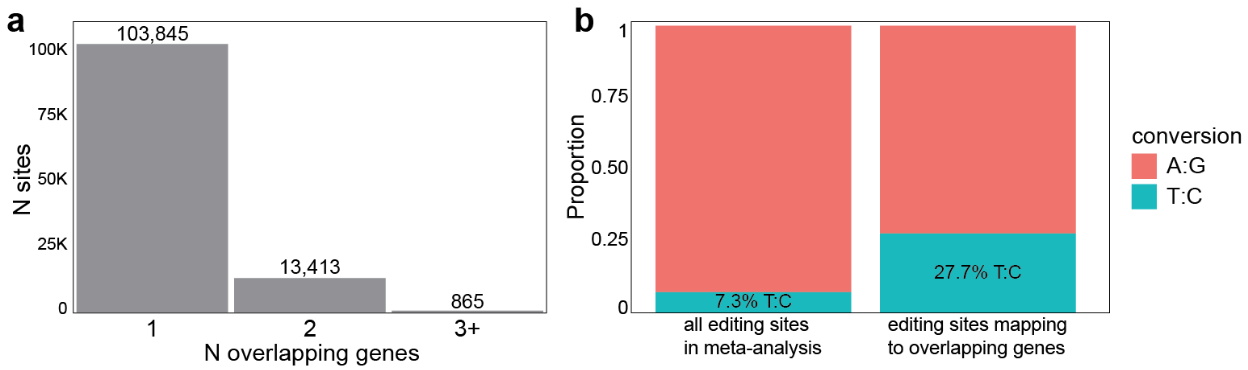


**Supplementary Figure 2 | Antisense A-to-I editing. a)** The number of editing sites which overlap multiple GENCODE v46 transcripts. **b)** Proportion of T:C conversions amongst all editing sites included in the meta-analysis versus those sites which overlap inversely oriented genes.

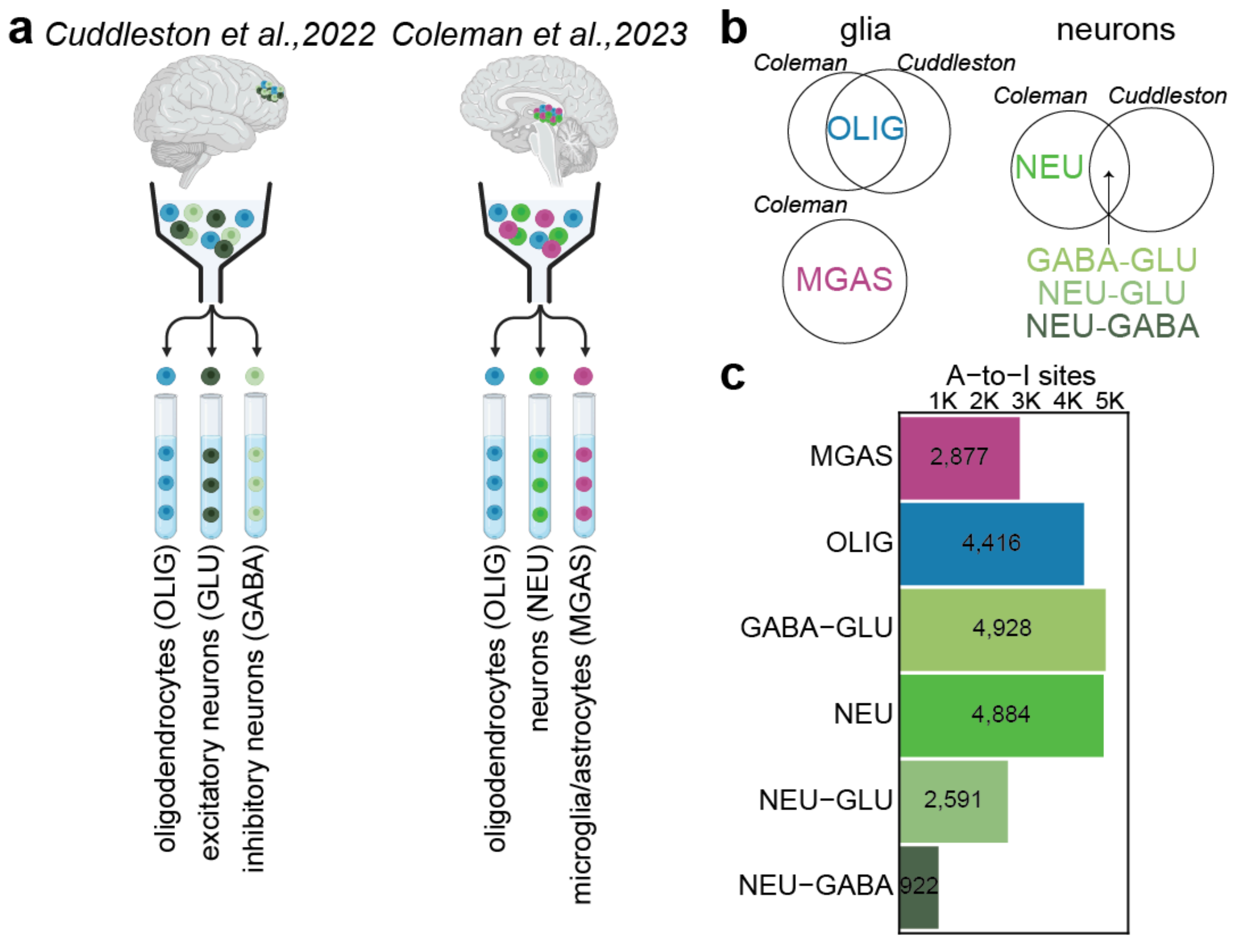


**Supplementary Figure 3 | Cellular specificity of A-to-I sites in BigBrain. a)** Depiction of the fluorescence-activated nuclei sorting (FANS) strategies from *Cuddleston et al., 2022* (*Cuddleston*) and *Coleman et al., 2023* (*Coleman*). In *Cuddleston*, three pools were sorted from prefrontal cortex tissue: oligodendrocytes ($NeuN^-/SOX10^+$), excitatory GABAergic neurons ($NeuN^+/SOX6^+$), and inhibitory glutamatergic neurons ($NeuN^+/SOX6^-$). In *Coleman*, three pools were sorted from parahippocampal gyrus tissue: oligodendrocytes ($NeuN^-/SOX10^+$), neurons ($NeuN^+/SOX10^-$), and pooled microglia/astrocytes ($NeuN^+/SOX10^+$). **b)** Editing sites were defined as cell-type specific within each study, and these categories were harmonized for cellular annotation of the sites identified in BigBrain (see Methods), where MGAS were distinct in *Coleman*; OLIG replicated in both studies; NEU were distinct in *Coleman*; GABA-GLU were sites shared by the GABA and GLU pools in *Cuddleston* which replicate in the NEU pool in *Coleman*; NEU-GLU and NEU-GABA were specific to their respective pools in *Cuddleston* and replicate in the NEU pool in *Coleman*. **c)** Of the 118,123 high-confidence A-to-I sites, the number of sites (x-axis) annotated to each cell type (y-axis) are represented by colored bars. MGAS: microglia/astrocytes; OLIG: oligodendrocytes; NEU: neurons; GLU: glutamatergic neurons; GABA: GABAergic neurons.

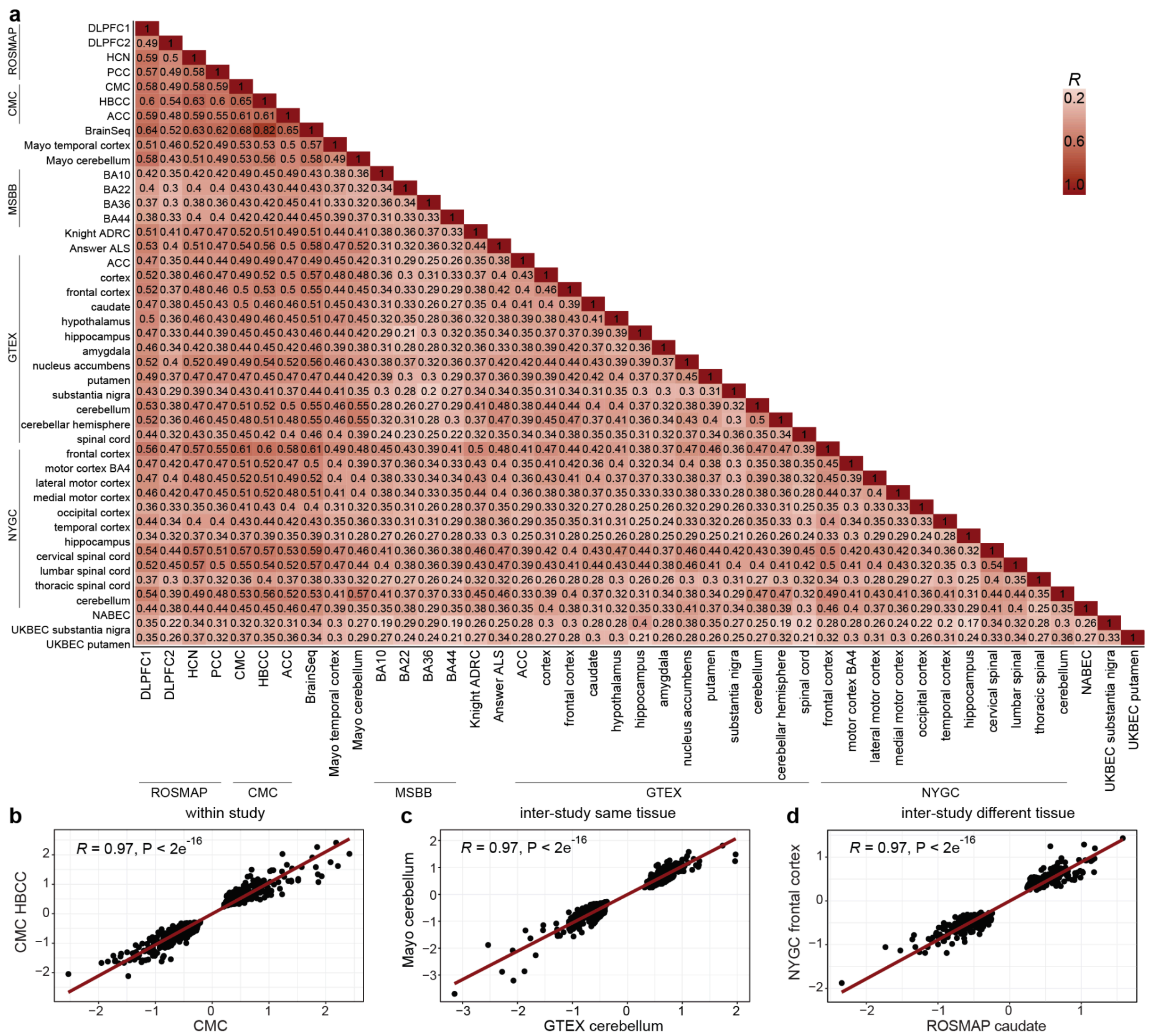


**Supplementary Figure 4 | Correlation of edQTL effect sizes between BigBrain datasets. a)** Pairwise correlation (Pearson) tests for all BigBrain datasets which were meta-analyzed. The heatmap is colored by the estimate (*R*), which is also displayed in each tile. Scatterplots highlight select correlations between two datasets which were from **b)** the same cohort, **c)** the same tissue across different cohorts, and **d)** different tissues from different cohorts, where the red trendline corresponds to the estimate and p-values displayed on each plot from a two-sided Pearson correlation test.

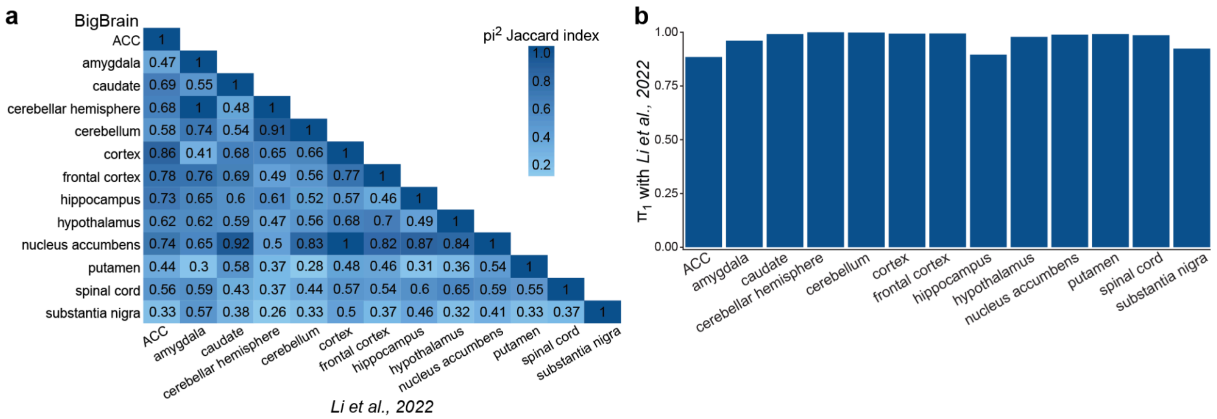


**Supplementary Figure 5 | Replication of previously published edQTLs in BigBrain. a)** $\pi^2$ analysis between all GTEx tissues from BigBrain (y-axis) and *Li et al., 2022* (x-axis). The heatmap is colored by Jaccard similarity index. **b)** Storey's $\pi_1$ replication (y-axis) of *Li et al., 2022* edQTLs in BigBrain for each GTEx tissue (x-axis).

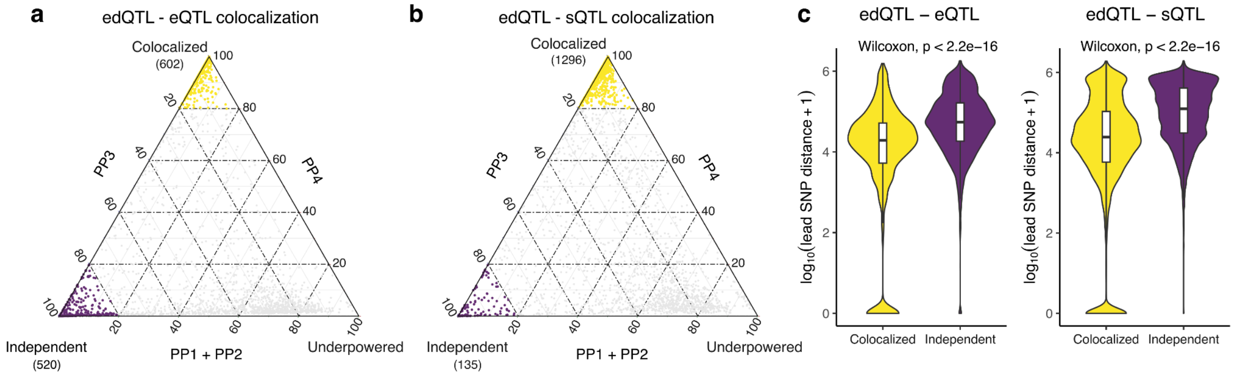


**Supplementary Figure 6 | Sharing of QTLs across editing, expression and splicing.** Ternary plots showing the posterior probabilities for pairs of **a)** edQTLs and eQTLs and **b)** edQTLs with sQTLs. PP3 is the posterior probability of the two QTL types being independent, and PP4 is the posterior probability of the two QTL types sharing the same single variant. **c)** Distance between the lead SNPs for pairs of QTLs.
